# Urinary cell-free DNA as a noninvasive liquid biopsy for hepatocellular carcinoma: a novel tool for diagnostics and monitoring

**DOI:** 10.1101/2021.12.03.21266943

**Authors:** Amy K. Kim, Selena Y. Lin, Hsin-Ni Liu, Surbhi Jain, Terence P. Gade, Fwu-Shan Shieh, Max Chao, James Hamilton, Hie-Won Hann, Ting-Tsung Chang, Dmitry Goryunov, Zhili Wang, Ying-Hsiu Su

## Abstract

**Background & Aims:** Cell-free DNA (cfDNA) has advanced cancer genetic profiling through liquid biopsy. While plasma is traditionally the primary source, emerging evidence highlights urinary cfDNA as a novel and noninvasive alternative. This study aimed to comprehensively assess transrenal DNA (trDNA) as a novel noninvasive biomarker source in HCC patients, compared to blood-based liquid biopsy.

**Approach & Results:** HBV DNA was used as a biomarker for trDNA. HBV-targeted and HCC-focused next generation sequencing (NGS) and whole genome sequencing (WGS) were used to compare fragment insert-sizes, the genome coverage, and germline genotyping accuracy. Urinary cfDNA overall exhibited a predominantly mononucelosomal pattern similar to plasma cfDNA, but with shorter fragments, broader size distribution and a more pronounced 10-bp periodicity. In contrast, trDNA were shorter and more variable among all patients. In HCC patients, trDNA was even shorter, with distinct 4-mer end motifs, compared to non-HCC trDNA. Higher concentrations of HCC-distinctive 4-mer end motif and TP53 mutations were found in urine compared to plasma. The overall genome coverage breadth by WGS was similar between urine and plasma cfDNA, with a higher fraction of covered cancer-associated mutation hotspots in urine cfDNA. In 101 HCC patients, there was a 78% overall concordance of HCC-associated mutations (*TP53, CTNNB1,* and *hTERT*) and in select 15 patients, 97% overall position-level concordance by targeted NGS between plasma and urine cfDNA.

**Conclusion:** Urine cfDNA has comparable features with distinct characteristics to plasma cfDNA and is a promising tool for liver cancer studies.

## Introduction

Cell-free tumor DNA (ctDNA) in biofluids has created new opportunities for cancer diagnostics and management. While most applications have focused on ctDNA in plasma, ctDNA from the bloodstream can filter through the kidneys into urine. Urine has been shown to contain ctDNA from both urological and non-urological cancers, including hepatocellular carcinoma (HCC) (1–18). This suggests that urine can serve as a noninvasive source of genetic and epigenetic information for HCC and as a potential alternative or complement to plasma for HCC liquid biopsy.

Urine provides a noninvasive approach that addresses the challenges of blood collection, such as limitations in specimen volume and frequency. Additionally, the convenience of at-home urine collection has the potential to enhance access to cancer screening, particularly in underserved populations (19).

Despite the promise of transrenal ctDNA, reports on its detection efficiency compared to blood ctDNA have been mixed, in both genitourinary (GU) (7, 20, 21), and non-GU cancers (14, 22–25). Urinary DNA is a complex mixture originating from prerenal, renal, and postrenal sources, it includes high-molecular-weight (HMW) DNA (>1,000 bp) from exfoliating urothelial cells and cell-free DNA (cfDNA) originating transrenally or from urinary tract cells (6, 18, 26). Unlike plasma, urine is not under stringent homeostatic regulation, and ctDNA detection is significantly affected by preanalytical factors and the assays used (27, 28). For instance, there is considerable variability in the reported sizes of transrenal DNA fragments, ranging from ∼50 bp to 150-300 bp, likely due to differences in isolation and assay platforms (29, 30). This variability complicates the assessment of the urine liquid biopsy utility.

This study evaluated urine liquid biopsy against plasma-based liquid biopsy in HCC. Using hepatitis B (HBV) DNA in urine samples from patients with chronic HBV infection (hepatitis), cirrhosis, and HCC, we analyzed transrenal DNA fragment size. Different sequencing techniques were employed to compare overall genome coverage, 4mer-end motifs, HCC hotspot coverage, and germline or somatic mutations concordance. Targeted NGS of matched plasma and urine further confirmed the detection of somatic variants. This approach offers a promising noninvasive alternative for genetic profiling, facilitating frequent sampling for early detection and disease monitoring of urine liquid biopsy in clinical practice.

## Materials and Methods

### Patients and body fluid collection

Detailed patient information is summarized in **Tables S1, S2,** and **S3**. Specimen collections were approved by each study center’s institutional review board. This study was conducted in accordance with both the Declarations of Helsinki and Istanbul. Urine was mixed with EDTA (10 mM final concentration) immediately upon collection and stored at −80°C until isolation. Blood was collected in K_2_EDTA BD Vacutainer tubes (BD, Franklin Lakes, NJ) and processed to obtain plasma and peripheral blood mononuclear cells (PBMC).

### DNA isolation, quantitation, and size distribution analysis

DNA was extracted from urine samples as previously described (31). The low molecular weight (LMW) fraction (cfDNA; < 1 kb) was size selected with HighPrep PCR magnetic beads (MagBio Genomics, Gaithersburg, MD) at 0.55X bead/sample ratio. Plasma cfDNA was extracted using the Quick-cfDNA Serum & Plasma kit (Zymo Research, Irvine, CA) or JBS plasma cell-free DNA isolation kit (Cat# 08874, JBS Science, Inc, Doylestown, PA). DNA concentrations were determined using the Qubit dsDNA HS assay kit (Thermo Fisher Scientific, Waltham, MA). DNA Size distribution was analyzed on Agilent’s 2200 TapeStation system using High Sensitivity D5000 screentapes (Agilent, Santa Clara, CA). PBMC DNA was isolated using the Genomic DNA Isolation Kit (Qiagen, Valencia, CA) according to the manufacturer’s instructions.

### Mutation detection by qPCR

Archived DNA from 101 HCC patients (15) was used for mutation testing. First, the total copy number of each target gene was determined for each DNA sample using quantitative PCR (qPCR) assays (JBS Science) according to manufacturer’s instructions. Only urine DNA samples that contained >100 copies/mL were used for analysis. Mutation detection in three markers of interest was then performed in duplicate using JBS mutation assay (JBS Science).

### NGS library preparation and sequencing and data analysis for HBV insert size and variants

For whole genome sequencing (WGS), fragmented DNA (LMW cfDNA or sonicated PBMC/total urine DNA) with DNA inputs ranged from 30 to 50 ng was subjected to Illumina NGS library preparation and sequenced on a NextSeq550 or HiSeq4000 instrument (2×150 bp; Genomics Core Facility at Drexel College of Medicine, Philadelphia, PA).

For targeted sequencing by JBS HBV or JBS HCC panel, libraries were prepared from 10 ng of urine or plasma cfDNA using the NEBNext Ultra II DNA Kit (NEB, Ipswich, MA) and xGen UDI-UMI adapters (IDT, Cornwall, IA) and subjected to enrichment according to the manufacturer’s protocol, and sequenced on a NovaSeq S1 flow cell (Psomagen, Rockville, MD) for paired-end sequencing. The HCC panel targets 24 genes with a panel size of 64 kb (**Table S4**).

Raw data were demultiplexed and each paired-end read was tagged with the UMI. Unmapped BAM (uBAM) files were then converted to fastq format for initial alignment using BWA MEM and hg19 as a reference. The resulting BAM files were downsampled within each plasma/urine pair. UMI processing was performed using fgbio, connor, and picard suites, as detailed in **Figure S1**. A minimum UMI family size of three was used in most analyses. Consensus reads were realigned, quality-filtered, and used to generate VCF files that were filtered for common germline variants (dbSNP138) and used to evaluate mutant variant frequencies.

## Results

### Urine cfDNA size distribution similar to plasma cfDNA

We performed capillary electrophoresis of LMW DNA isolated from urine samples of eight healthy individuals and eight HCC patients. As anticipated, mononucleosomal DNA (150-180 bp) was the predominant cfDNA species in all samples (**Figure 1**). Most samples also contained a minor dinucleosomal peak or “shoulder”.

**Figure 1.**
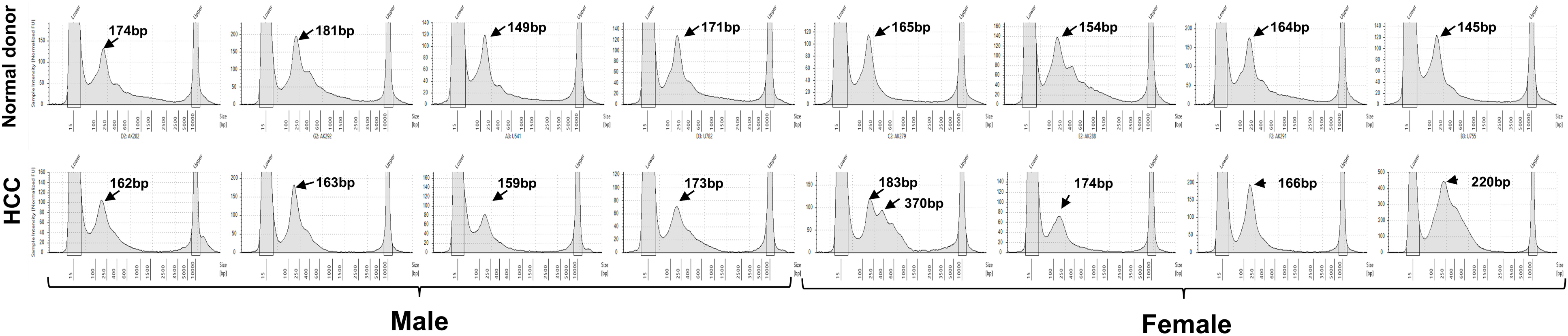
Electrophoretic size profiling of LMW DNA isolated from urine. Eight healthy and eight HCC urine LMW DNA samples were resolved on a TapeStation as described in Materials in Methods. Healthy (top) and HCC (bottom) sample panels are indicated. Equivalents of approximately 0.1 mL of healthy-donor urine and 0.057 mL of HCC-patient urine were loaded.

Next, shallow whole genome sequencing (sWGS) of LMW DNA isolated from three sets of matched urine and plasma samples from two patients with HCC (**Table S1,** LMW DNA profiles shown in **Figure S2**) was performed. Sample ID649 and ID697 were collected from the same patient eight months apart. Samples 649 from both urine and plasma were also resequenced at a higher depth. The WGS data are summarized in **Table S5**.

All plasma samples showed a major peak at 170-180 bp, corresponding to the mononucleosomal fragment size (**Figure 2A-D**) with a minor dinucleosomal peak. Urine cfDNA showed a similar mononucleosomal peak, but no dinucleosomal peaks were discernable. Both plasma and urine insert size distributions exhibited a subnucleosomal “sawtooth” pattern with a 10 bp periodicity, more pronounced urine. Consistent with this observation, the insert size distribution in the urine samples was wider compared to that in the plasma libraries (**Figure 2E**). Sequencing at a higher depth revealed no notable differences in fragment size distributions in both urine and plasma compared to sWGS.

**Figure 2.**
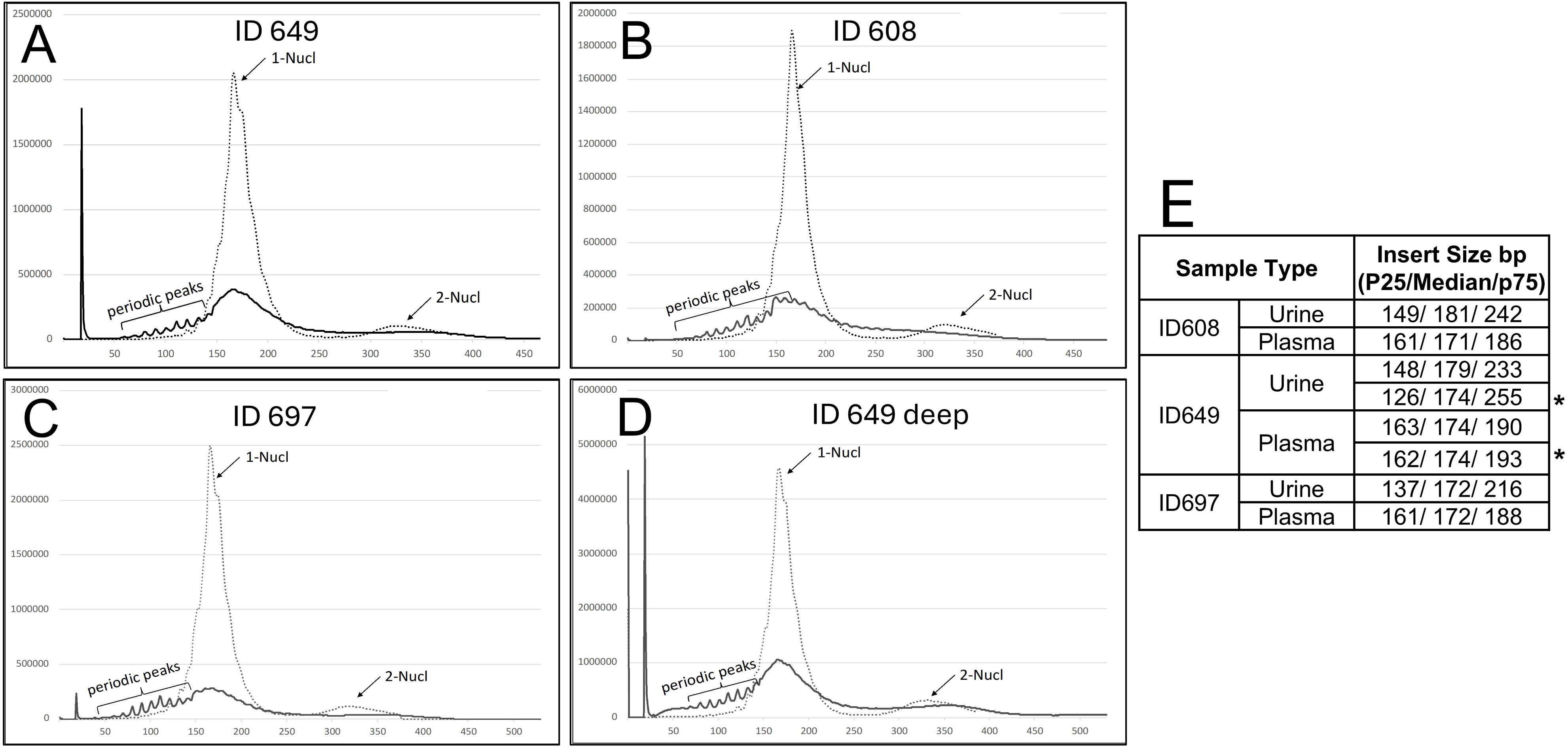
Analysis of WGS insert size distributions in matched urine and plasma cfDNA samples from patients with HCC. (A-C) Insert size distributions in shallow-sequenced samples. (D) Samples 649 resequenced at higher depths. Plasma and urine samples are dashed and solid lines, respectively. Mono- (1-Nucl) and di- (2-Nucl) nucleosomal peaks are indicated. The 10 bp periodic peaks are indicated with brackets. (E) Median and 25^th^/75^th^ percentile insert sizes for each sample. In ID649, the bottom urine/plasma rows (*) are for the deeper WGS run. See **Table S5** for details.

### Urinary HBV DNA is significantly shorter than human cfDNA in urine

Urinary HBV DNA which is liver-derived was used as a biomarker to examine the size of transrenal DNA (trDNA). Human cfDNA in urine (blue, **Figure 3A**) exerted a mono-nucleosomal peak with pronounced a subnucleosomal “sawtooth” pattern with 10 bp periodicity, similar to the earlier size distribution pattern (**Figures 1 & 2**).

**Figure 3.**
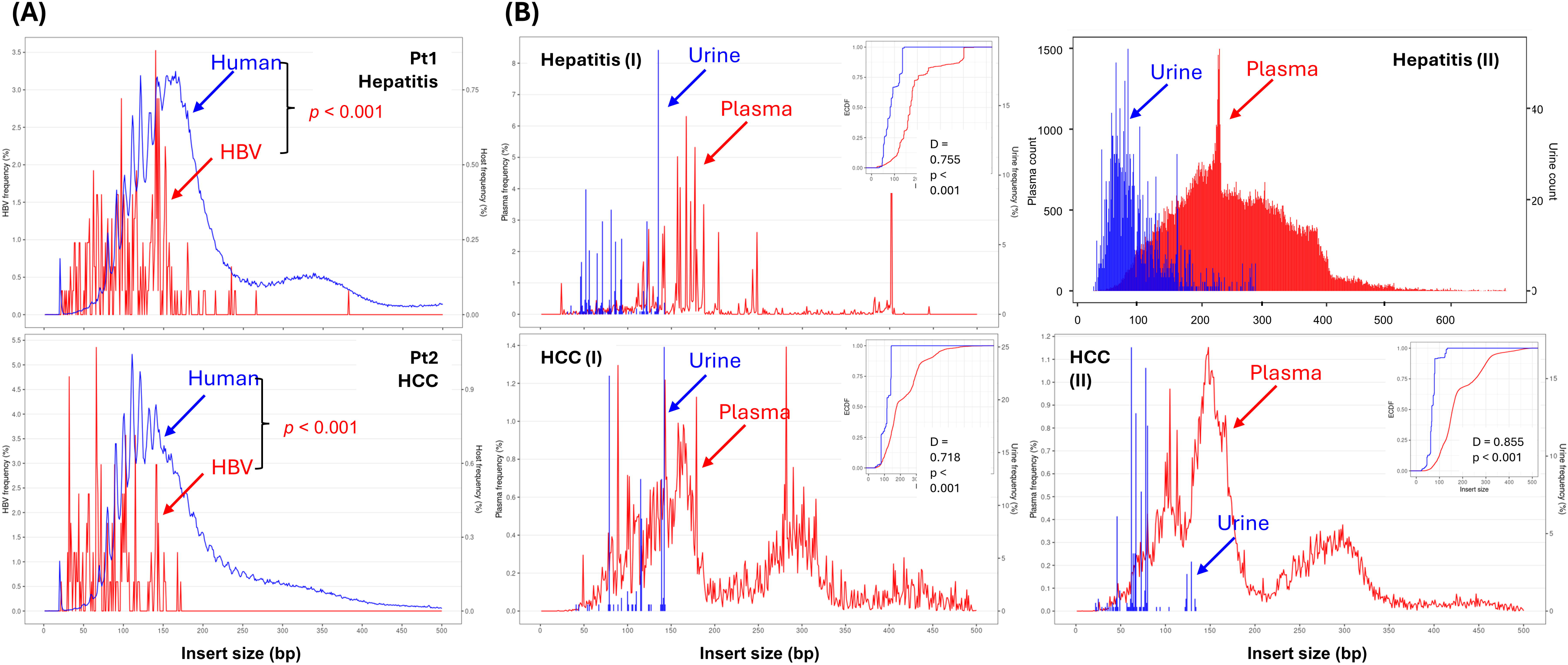
trDNA size distribution as compared to human cfDNA in urine (A) and liver-derived cfDNA in plasma (B). (A) Size distribution between trDNA (HBV DNA, red) and urine cfDNA (human, blue), as indicated from one patient with HBV hepatitis, Pt 1 and one HBV-associated HCC patient, Pt 2. (B) HBV DNA size distribution between matched urine and plasma from two hepatitis B (I & II) and two HCC (I & II) patients. Note, HBV hepatitis (II) patient had a 6.9 log IU/mL serum viral load at the time of specimen collection.

We compared HBV DNA insert sizes (red, **Figure 3A**) which represent fewer reads than human cfDNA, in urine samples from two patients: Pt 1 with HBV and Pt 2, a patient with HBV-associated HCC. The HBV DNA is significantly shorter than human cfDNA in urine (p<0.001, by student t-test) for both examples.

Next, we compared the HBV DNA size distribution between urine and plasma from individuals collected on the same day to control for temporal variations and technical variables (**Figure 3B**). To avoid possible contamination from viral particles in the blood, we selected matched urine and plasma from three patients with undetectable serum viral load, Hepatitis (I) and HCC (I and II). A hepatitis B patient with high viral load, 6.9 log IU/mL, Hepatitis (II) was included as a control. As expected, plasma HBV DNA showed typical nucleosomal sizes, whereas HBV trDNA showed significantly shorter fragment sizes, peaking in a wider range of 80-150 bp which suggests greater individual variation (**Figure 3B**). By examining urine and plasma from a high viral load patient Hepatitis (II), trDNA size distribution was similar to that of patients with undetectable viral load, Hepatitis (I) and HCC (I & II). The size of circulating HBV DNA in plasma of the patient with high viral load (Hepatitis II) was less focused in nucleosomal sizes and peaked at 228 bp in this individual, possibly derived from circulating viral particles.

### Comparison of HBV DNA insert size among three disease groups, hepatitis, cirrhosis, and HCC

It has been demonstrated that ctDNA is shorter than cfDNA in the blood (32). Since trDNA in urine is derived from blood, we hypothesized that shorter urinary HBV DNA would be observed in patients with liver cancer. We thus compared HBV DNA size distribution in urine from patients with HCC (n=17) to those with hepatitis (n=14) and cirrhosis (n=13) (**Figure 4A-C)**. As expected, urine HBV DNA differed significant among the three disease groups (p<0.001, one way ANOVA) as summarized in **Figure 4D**. HBV DNA in HCC patients was the shortest (cirrhosis-HBV: p<0.001, cirrhosis-HCC: p=0.048, HBV-HCC: p<0.001, pair-wise comparison).

**Figure 4.**
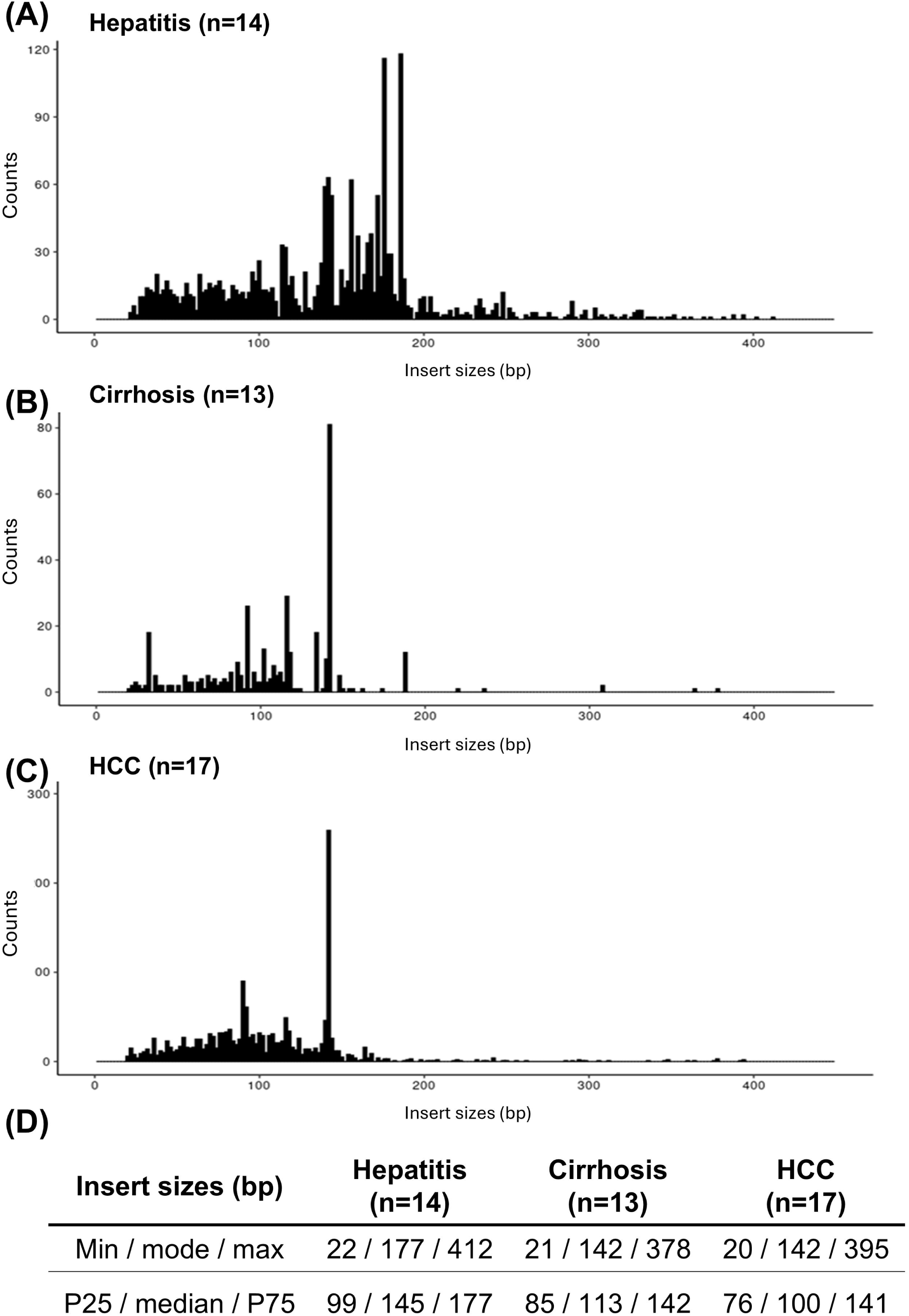
Comparison of HBV DNA fragment size in urine from 14 hepatitis, 13 cirrhosis or 17 HCC patients. Insert sizes of the HBV DNA collectively from three disease groups, hepatitis (A), cirrhosis (B), and HCC (C) are plotted by NGS read counts per size. Panel D compares the median with P25 and P75 from each disease group.

### Comparison for 4-mer end motifs of HBV DNA between patients with or without HCC in urine and plasma

cfDNA fragmentation has been shown to be nonrandom with various DNA nucleases generating sequence-specific cleavage or unique 5’ end motifs. These end motifs have been found to distinguish between cancer and non-cancer patient plasma samples including those with HCC (33–36). Hence, we analyzed the 4-mer end motifs of HBV DNA between HCC and non-HCC HBV patients to determine whether distinct signatures can also be observed in trDNA and whether these signatures are similar between urine and plasma.

We compared the motifs frequency of the 10 most frequent HBV DNA 4-mer end motifs in plasma (**Figure 5A**, top panel) and in urine (**Figure 5B**, top panel) from HBV patients with HCC (HCC, n=17, blue) and without HCC (CHB, n=14, red) (**Figure 5**, bottom panels). Overall, there is only one, CGTT, overlapped between the 10 most frequent motifs identified in plasma and urine. CGTT is the 10^th^ most frequent observed motif in plasma and the 3^rd^ most frequent motif in urine, but for both plasma and urine, frequency was much lower in HCC than non-HCC. Although the first 9 most frequent motifs in plasma did not exert detectable difference between HCC and non-HCC, the first two, CCCA and CCAA, in urine were found to be much more abundant in HCC than in non-HCC. Specifically, in plasma, CCAA, had no appreciable difference in frequency between patients with and without HCC, but found to be three times more abundant in urine from HCC patients compared to non-HCC. Similarly, the second most frequent 4-mer end motifs in urine, GAGA, was 6-fold more in HCC urine than in non-HCC urine, but no detectable difference found in plasma. Note, GAGA was not part of the 10 most frequent 4-mer end motifs identified in plasma. Collectively, the comparison of these 4-mer motifs suggests the difference between HCC and non-HCC is more pronounced in trDNA compared to cfDNA in plasma.

**Figure 5.**
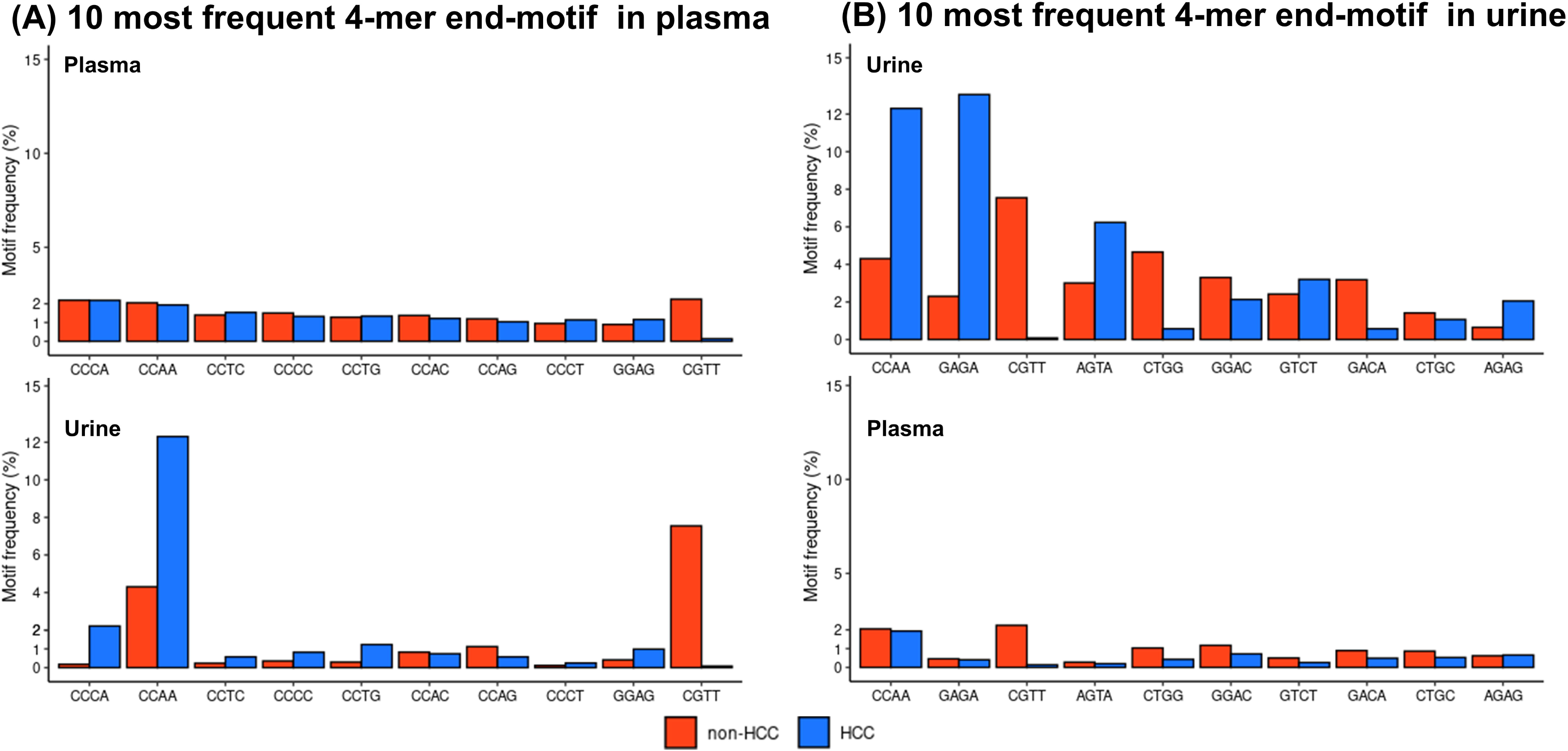
Comparison of the 10 most frequent 4-mer end motifs abundance in HBV DNA fragments in plasma (A) and in urine (B, top panel) between HCC (blue) and non-HCC HBV hepatitis (red) patients. The 10 most frequent 4-mer end motifs identified in plasma (A) and in urine (B) listed by the motif frequency (%)(top panel) and corresponding matched plasma or urine (bottom panel) from HCC (blue) and non-HCC (HBV hepatitis, red) patients are compared.

### Comparison of genome representations in urinary and plasma cfDNA from patients with HCC

The total fractions of overall genomic positions covered at least once in urine and plasma cfDNA were determined and demonstrated similarity within each pair as detailed in **Table S5**. Overall, 87.5% to 94.4% of the urine cfDNA coverage overlapped with the genomic positions covered in the matched plasma cfDNA. To examine the difference in sWGS genome coverage between matched plasma and urine cfDNA in more detail, we visualized the overlap of genomic positions covered in each pair using Euler diagrams (**Figure 6**). The urine/plasma overlap ranged from 48% to 58% of the genome in the three shallow-sequenced sample pairs and increased to 60% to 73% with a higher depth sequencing of samples (pt.649). Positions covered in plasma, but not in urine comprised 6-8% of the genome, whereas the fraction of urine-only coverage was 7-13% across all pairs. Moreover, despite the cfDNA inputs of the plasma libraries being 2- to 8-fold higher than those of the urine, comparable overall coverage and high coverage overlap between plasma and urine were observed.

**Figure 6.**
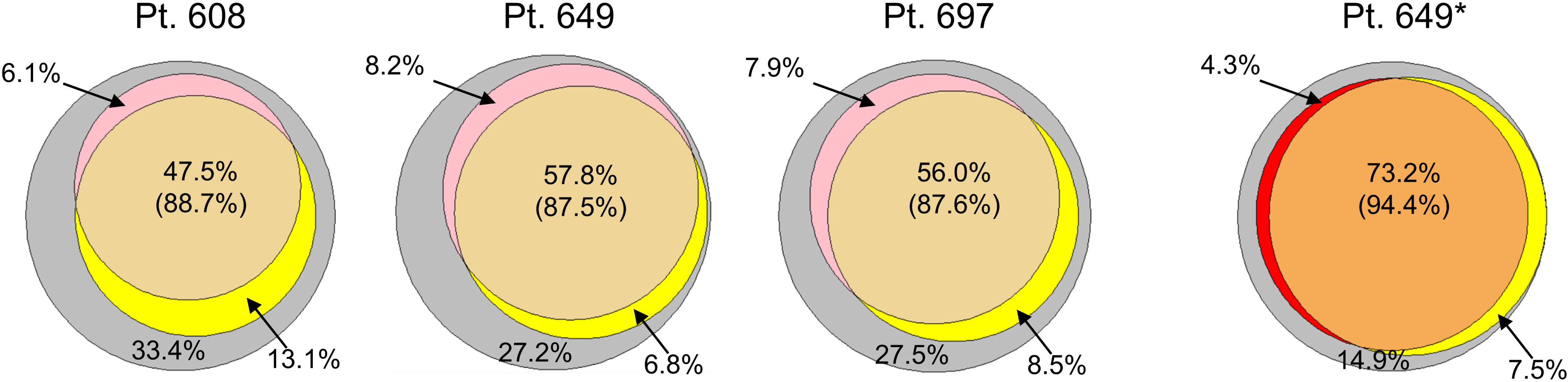
Genomic coverage of LMW cfDNA isolated from matched urine (yellow) and plasma (pink/red). Euler diagram of genomic positions in urine and plasma covered in each pair. The urine/plasma overlap ranged from 48% to 58% of the genome in the three shallow-sequenced sample pairs and increased from 60% to 73% when the ID649 samples were sequenced to higher depths. Positions covered in plasma, but not in urine comprised 6-8% of the genome (pink area), whereas the fraction of urine-only coverage was 7-13% across all pairs (yellow). * re-sequenced to higher depth. Grey indicates the genomic sequence not covered by urine or plasma.

Next, we compared cancer hotspot coverage obtained in the two matched sample types using the downsampled sWGS data. The hotspot regions analyzed corresponded to the NEBNext™ Direct hotspot cancer panel which is comprised of 190 target regions in 50 genes (37). The hotspot coverage in urine was comparable to, or slightly higher than, the corresponding plasma coverage in each pair as detailed **Figure S3A**. In deep-sequenced pair of ID649 samples, both plasma and urine cfDNA covered approximately 94.5% of the hotspot target positions. These findings are consistent with previous reports of cancer-associated mutations being enriched in urine DNA in other solid tumors and our earlier work (5, 16, 38).

We then visualized the hotspot coverage in all three sample pairs using Circos plots (**Figure S3B and C**). Although the coverage profiles were divergent between plasma and urine, they were similar within each sample type. This observation suggests that while the genome may be represented differentially in plasma and urine cfDNA, the biases are qualitatively reproducible within sample types. This finding was further supported by hotspot coverage profiles in deep-sequenced sample pair ID649 being similar to those seen in the shallow-sequenced counterparts (data not shown).

### TP53, CTNNB1, and TERT mutation prevalence in matched urine and plasma cfDNA from patients with HCC by qPCR

Because the sWGS results showed differential genomic coverage in urine and plasma cfDNA, we compared the prevalence of HCC-associated mutations between the two sources of ctDNA. HCC is most frequently linked to mutations in the *TERT*, *TP53*, and *CTNNB1* genes (39, 40). We have previously developed short-amplicon qPCR assays for the detection of mutations in the *TERT* promoter (c.-124), *TP53* (codon 249), and *CTNNB1* (codons 32-37) (4, 16) with sensitivities at 0.15%, 0.1%, and 0.3%, respectively. Urine and plasma cfDNA samples isolated from a total of 101 patients with HCC were screened. The results are detailed in **Table S6** and summarized in **Table 1**.

**Table 1.**
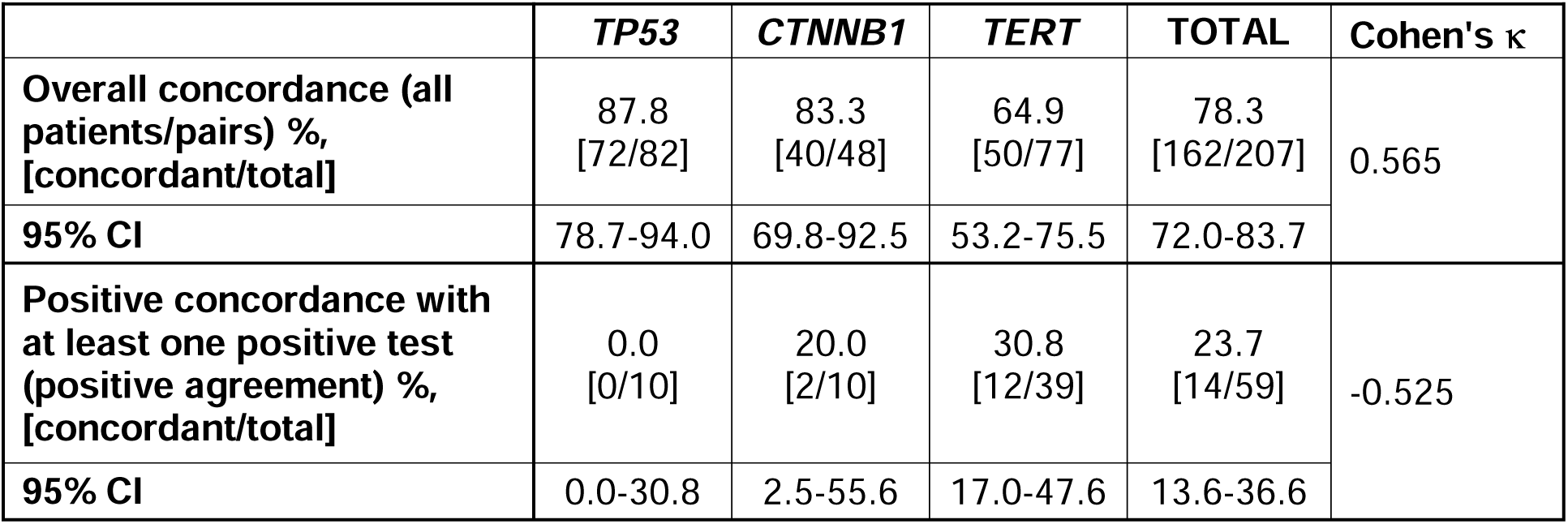
Detection of ctDNA markers by qPCR in matched urine and plasma samples from patients with HCC (n=101).

The difference in overall non-matched detection rate was not statistically significant between plasma and urine (18.4% vs. 16.9%; p=0.797, Fisher’s exact test). The overall concordance (i.e. across all patients, target genes, and urine-plasma pairs, including those with double negative results) was 78%. However, the number dropped to 24% when only urine-plasma pairs with at least one positive result were counted (positive agreement) among all target genes. In this group, *TERT* showed the highest concordance (31%), followed by *CTNNB1* (20%) and *TP53* (0%). Notably, *TP53* mutations were detected more in urine than in plasma (p=0.023; Fisher’s exact test). No significant differences in *CTNNB1* or *TERT* mutation prevalence were observed between plasma and urine within pairs with at least one positive result (**Table S7**).

### Targeted NGS analyses of urine and plasma cfDNA from HCC patients

Next, we performed hybridization-based target enrichment followed by NGS to evaluate the potential of utilizing matched urine and plasma cfDNA for somatic mutation detection in 15 patients with HCC (**Table S3**). All data were downsampled to approximately 2.2×10^7^ paired-end reads for comparative analysis and consolidated by UMI with a minimum family size of three, resulting in mean per-base on-target coverage of approximately 1,500× and 1,280× for plasma and urine, respectively (**Table S8**). The global (i.e. across all patients/target positions) variant call rates were significantly higher in urine compared to plasma (0.0266 vs. 0.0186; McNemar χ^2^=1,942.6; p<0.0001), potentially indicating either a higher background noise in the urine data, urine being enriched for ctDNA relative to plasma, or both. The global position-level concordance was 96.8%, while the both-variant positive agreement was 17.7%. The concordances for plasma- and urine-variant positive positions were 36.5% and 25.5%, respectively (**Table S9**). The global mean variant allele frequencies (VAFs) were similar between the two body fluids, albeit the difference was significant (0.046% for plasma vs. 0.048% for urine; Wilcoxon signed-rank test p<0.0001). Consistent with this result, plasma and urine VAFs for double-positive positions showed strong global correlation (Pearson r=0.966 [95% CI 0.966-0.966]; **Figure S4A**). The correlation was even stronger for position-level mean VAFs across the entire cohort (Pearson r=0.995 [95% CI 0.995-0.995]; **Figure S4B**). As expected, putative somatic VAFs (<0.4) correlated less strongly (Pearson r=0.890 [95% CI 0.884-0.895]; **Figure S4C**).

Among the 24 target genes, *TERT* promoter showed the highest somatic variant call rate in both plasma and urine with 20.2% and 21.9%, respectively, followed by *KMT2D* (3.7 and 4.2%), *TP53* (2.8 and 3.5%), and *ARID1A* (2.4 and 3.5%). Mutation rates were overall similar between urine and plasma, but in six targets, urine had more than a two-fold higher variant rate than in plasma (**Table S10**). Putative somatic mutations with 100% sample-level (n=30) frequencies were found in *ARID1, hTERT* promoter, *PCLO, KMT2D, AXIN1, TSC2, TP53*, and *KEAP1* (**Supplemental File 1**). Taken together, these results suggest that the mutational profiles of HCC tumors are likely to be reflected similarly in urine and plasma cfDNA.

### Comparison of blood- and urine-derived DNA as a source of germline genotype information

Liquid biopsy assays using blood as the source of genetic material often rely on sequencing DNA extracted from PBMCs to determine the patient’s germline genotype. In addition to cfDNA, urine contains exfoliated urothelial cells as well as some leukocytes (41), which normally contain the individual’s germline genetic information. For a urine-based liquid biopsy assay to be self-sufficient, both somatic and germline genotype information needs to be recoverable from urine DNA. To demonstrate the feasibility of germline genotyping using total urine DNA, we first performed WGS of PBMC/total urine DNA pairs isolated from six healthy subjects (**Table S11**). In all pairs, the genome fraction covered in urine DNA was ≥ 99.3% relative to the matched PBMC coverage. Next, we evaluated genotype concordance across all positions sequenced at a minimum depth of 10 in both sample types. Perfect concordance was observed in all six sample pairs (**Table 2**). These results demonstrate that total urinary DNA can be used for germline genotyping similarly to DNA isolated from PBMCs.

**Table 2.** Germline genotype concordance between PBMC and total urine DNA.

| **ID** | **Number of bases covered at least 10X in both DNA samples** | **Number of concordant bases** | **Concordance** |
| --- | --- | --- | --- |
| **#1** | 2,455,224,425 | 2,455,224,425 | 100% |
| **#2** | 2,348,155,979 | 2,348,155,979 | 100% |
| **#3** | 2,443,568,212 | 2,443,568,212 | 100% |
| **#4** | 2,417,859,203 | 2,417,859,203 | 100% |
| **#5** | 2,120,546,181 | 2,120,546,181 | 100% |
| **#6** | 2,365,129,319 | 2,365,129,319 | 100% |

## Discussion

This study demonstrated that urinary cfDNA provides a viable, noninvasive source for genetic profiling, capable of detecting both germline and somatic mutations. We analyzed genomic features including DNA fragment size distribution, 4-mer end motifs, HCC-associated mutation detection, and genome coverage of trDNA in comparison to plasma cfDNA to evaluate the potential of urine-based liquid biopsy as a noninvasive alternative or complement to blood-based liquid biopsy for HCC. Notably, based on HCC characteristic 4-mer end motifs, *TP53* mutations, and the HCC-focused NGS (**Table S8**) VAF analysis, urine may contain a higher concentration of some HCC ctDNA. By WGS, cfDNA from urine and plasma have similar overall genome coverage with mostly overlapped positions. HCC-associated mutations were also detectable at similar rates overall between urine and plasma cfDNA by qPCR and targeted NGS which revealed high concordance between sequence variants detectability in the two body fluids.

We previously demonstrated that HBV DNA detected in urine originates from chronically infected hepatocytes unless there is blood contamination. Using HBV DNA as a trDNA marker to compare trDNA with total cfDNA in urine, and samples between HCC and non-HCC patients, several notable characteristics were found. **First**, HBV trDNA from non-cancer patients is shorter and has a broader range in its size distribution than plasma cfDNA, with more variance in size peaking at ∼50 bp, 80 bp, 100 bp or even longer. Our analysis was from all DNA isolated without pre-processing centrifugation to avoid losing cell debris associated trDNA (31, 42). This may explain discrepancies for the trDNA size reported in the literature (43) ranging from mostly 50 bp or less, with median sizes of 80 or 100 bp (29, 30). **Second**, trDNA is shorter in HCC than non-HCC, which is consistent with previous studies in blood. **Third**, the most frequent 4-mer end motif, CCCA, which was previously identified in plasma cfDNA, is also the most frequent motif identified in HBV DNA in plasma, although the abundance of HBV CCCA motif shows no detectable difference in plasma between HCC and non-HCC, as shown in a previous study (33). Although CCAA is not among the 10 most frequent motifs observed in urine, it was significantly more abundant in HCC patients compared to non-HCC patients. This finding contrasts with previous plasma-based studies, where CCAA was observed to be less abundant in HCC patients compared to non-HCC patients. Other studies do not show a significant difference in CCAA between cancers and controls as analyzed by cfOmics (44, 45). While there may be some inconsistency in 4-mer motifs comparison between studies, our data using HBV DNA motifs suggests the end motifs frequencies in urine may show better distinction between HCC and non-HCC, compared to plasma. This is consistent even in the detection of TP53 mutation, where significantly more TP53 mutations were detected in urine than in plasma. This is similar to findings in urothelial cancers (46, 47) and NSCLC (25).

Using mutation-specific qPCR assays, we found a high overall concordance of 78% in 101 patients screened for three HCC hotspots mutations and the overall non-matched detection rates were not significantly different between plasma and urine. Positive agreement between plasma and urine was only 24%, suggesting that ctDNA representation may differ between urine and plasma even in patients with non-genitourinary malignancies (48). This is in contrast with other plasma-urine *EGFR* T790M positive agreement reports in other cancers such as NSCLC (11, 38) and bladder (49). The apparent discrepancies in urine/plasma positive agreement between these studies and our results may be attributable to the pre-analytic variables such as detection assays and DNA isolation method and fragmentation extent of different genes. Similar reasons may underlie the relatively low plasma-urine positive agreement (18%) in the targeted NGS data from additional 15 patients with HCC. It is also important to note that most patients in our cohort had early-stage disease and our library inputs were only 10 ng, both of which may have substantially reduced the limit of detection sensitivity in our study. Tumor tissue mutational profiles were unavailable for most patients, limiting our ability to analyze any concordance between tissue and plasma/urine DNA. However, it is well-known that mutation profiles often differ between tissue samples and plasma. This discrepancy arises because tissue analysis typically relies on fine needle biopsies which is only a very small fraction of the highly heterogenous tumor nodule(s) whereas plasma cfDNA represents a collection of cfDNA from the dying cells from the entire tumor (50).

Lastly, urine was also determined to be a reliable source of germline genotype information, similar to PBMCs in blood-based liquid biopsies with high genome fraction coverage. Perfect genome coverage concordance was obtained between urine DNA and the matched PBMC. Thus, similar to PBMC-derived DNA in blood-based liquid biopsy, total urinary DNA can be used to extract germline information.

Our study has several limitations. First, the disparate quantities of plasma and urine cfDNA used for WGS (but not targeted NGS) hindered a rigorous comparison of some WGS metrics and underlying biological phenomena. Second, due to the limited DNA available, the number of HCC patients analyzed by NGS was relatively small. Third, the DNA isolation method we used to obtain urinary cfDNA may not allow for efficient recovery of fragments <100 bp, which may be enriched for tumor-derived somatic variants (51). Lastly, the validity of comparative analyses of many liquid biopsy studies is greatly limited by the lack of a standardized approach to both DNA isolation and data acquisition technologies. Even within the same methodology, seemingly minor differences (e.g., sequencing depth or input amounts in the case of NGS) can affect study outcomes and conclusions drastically. It is therefore critical to reproduce our findings in larger-scale studies.

In conclusion, our results highlight urinary cfDNA’s potential as a noninvasive tool for liver cancer diagnostics, monitoring, and personalized treatment strategies. It offers a promising complement—or even alternative—to plasma-based approaches. The ability to reliably detect key genomic features and mutations underscores its clinical utility, while the ease of urine collection and potential for improved patient compliance make it particularly attractive for broader application in liquid biopsy. By addressing challenges associated with blood-based methods, urinary cfDNA could play a transformative role in early detection, disease monitoring, and access to personalized care in hepatocellular carcinoma. Future studies will be essential to validate these findings and further optimize this approach for widespread clinical adoption.

## Data Availability

All data produced in the present study are available upon reasonable request to the authors

## Author Contributions

A.K. obtained patient specimens and co-wrote the manuscript; S.L. devised the experimental plans, supervised execution, and analyzed the data; S.J. conceived and executed the experiments; F.S.S., H.L., and M.C. performed data analysis and produced figures; Z.W. developed cfDNA isolation for the study. D.G. performed the data analysis and co-wrote the manuscript; T.G., J.H., H.W.H., and T-T.C. obtained patient specimens; Y.H.S. conceived the study, coordinated the experimental plans, and co-wrote the manuscript.

## List of Abbreviations

cfDNA: cell-free DNA
ctDNA: cell-free tumor DNA
GU: genitourinary
HBV: hepatitis B virus
HCC: hepatocellular carcinoma
HMW: high-molecular-weight
LMW: low molecular weight
NGS: next generation sequencing
PBMC: peripheral blood mononuclear cells
qPCR: quantitative PCR
sWGS: shallow whole genome sequencing
trDNA: transrenal DNA
UMI: unique molecular identifier
VAFs: variant allele frequencies
WGS: whole genome sequencing

