## Supplemental data for "Urinary cell-free DNA as a noninvasive liquid biopsy for hepatocellular carcinoma: a novel tool for diagnostics and monitoring"

**Author’s affiliations:**

^1^Division of Gastroenterology and Hepatology, Johns Hopkins School of Medicine. Baltimore, MD 21205

^2^JBS Science Inc., Doylestown, PA 18902

^3^Baruch S. Blumberg Institute, Doylestown, PA 18902

^4^Department of Radiology, University of Pennsylvania Hospital, Philadelphia, PA 19104

^5^Division of Gastroenterology and Hepatology, Thomas Jefferson University Hospital, Philadelphia, PA 19107

**Corresponding Author:**

*Ying-Hsiu Su, The Baruch S. Blumberg Research Institute, 3805 Old Easton Rd, Doylestown, PA, 18902; Tel: 215-489-4949; Fax: 215-489-4920;.

**SUPPLEMENTAL FIGURES AND TABLES**

[**Table S10.** Somatic (VAF<0.4) variant call rates by target in the 15-patient HCC cohort (≥3 supporting UMI families).	13](#_Toc186808684)

### **Table S1.** Characteristics of HCC patients analyzed by whole-genome NGS.

| **Sample ID** | **Collection #** | **Sex** | **Gender (M/F)** | **Serum AFP (ng/ml)** | **Old case of HCC** | **Previous treatment dates and notes** | **HBV** | **HCV** |
| --- | --- | --- | --- | --- | --- | --- | --- | --- |
| 608 | 1 | F | 66-70 | 2.2 | 2010 | TACE in 11/2010 and 6/2015, new lesion 8/31/16 | - | + |
| 649 | 1 | M | 66-70 | 2.8 | 2004 and 2014 | TACE+RFA in 6/19/04, TACE in 04/14/14, MRI shows 0.9 cm LIRADS 4 recurrence on 3/2017 and 1.9 cm on 06/23/17 | + | - |
| 697  (Same patient as 649) | 2 |  | 66-70 | 4.2 |  |  |  |  |

### **Table S2.** Clinical information of HBV patients.

|  | **Hepatitis**  **(n=14)** | **Cirrhosis (n=13)** | **HCC**  **(n=17)** |
| --- | --- | --- | --- |
| Age (years) | 53.2  (35-70) | 58.5  (46-72) | 65.1  (42-82) |
| Gender (M:F) | 11:3 | 10:3 | 12:5 |
| Serum HBV DNA (log IU/ml)  <1.0 or UD  1-10  >10-10^2^  >10^2^-10^3^  >10^3^  NA | 14  0  0  0  0  0 | 11  0  1  0  0  1 | 4  0  2  3  5  3 |

UD, undetectable; NA, not available

### **Table S3.** Clinical information of 15 HCC patients for HCC-targeted NGS study.

| **Patient ID** | **Age** | **Gender (M/F)** | **Serum AFP (ng/ml)** | **HBV (+/-)** | **HCV (+/-)** | **Cirrhosis (Yes/No)** | **Alcoholic (Yes/No)** | **Primary or recurrent HCC** | **Stage of tumor (BCLC)** | **Tumor size (cm)** |
| --- | --- | --- | --- | --- | --- | --- | --- | --- | --- | --- |
| HCC-008 | 61-65 | M | 4.5 | - | + | No | No | Recurrent | B | 2.6 |
| HCC-030 | 56-60 | M | n/a | - | + | Yes | No | Primary | B | 2.3 |
| HCC-054 | 66-70 | F | 5 | - | - | Yes | No | Primary | C | 0.9 |
| HCC-057 | 61-65 | M | n/a | - | + | Yes | No | Primary | B | 2 |
| HCC-058 | 71-75 | M | n/a | - | + | Yes | Yes | Primary | B | 2 |
| HCC-060 | 66-70 | F | 5 | + | - | Yes | No | Recurrent | B | 2.9 |
| HCC-063 | 66-70 | M | 2.8 | - | - | Yes | No | Recurrent | B | 2.3 |
| HCC-068 | 66-70 | M | 5.6 | - | - | Yes | Yes | Primary | B | 2.2 |
| HCC-069 | 61-65 | F | 5 | - | + | Yes | No | Recurrent | A | 2 |
| HCC-079 | 76-80 | M | 8.3 | - | - | No | No | Primary | A | 2.5 |
| HCC-277 | 56-60 | M | 6 | - | - | Yes | No | Primary | A | 1.8 |
| HCC-297 | 61-65 | M | 5.3 | - | - | Yes | No | Primary | B | 4.4 |
| HCC-326 | 51-55 | M | 9.5 | - | - | Yes | Yes | Primary | A | 2.8 |
| HCC-335 | 66-70 | M | 7.5 | - | - | Yes | No | Primary | B | 2.6 |
| HCC-724 | 66-70 | M | 3.4 | - | n/a | n/a | n/a | Recurrent | n/a | 3.6 |

n/a, not available

### **Table S4.** 24 gene targets of JBS HCC NGS panel.

**Gene name**

ARID1A

APOB

LRP1B

NFE2L2

CTNNB1

BAP1

PIK3CA

ALB

TERT_promoter

IL6ST

APC

ARID1B

PCLO

CDKN2A

ATM

ARID2

KMT2D

HNF1A

RB1

AXIN1

TSC2

TP53

KEAP1

RPS6KA3

### **Table S5.** Summary of WGS data.

| **Sample**  **ID / Type** | | **Input, ng*** | **Total Reads** | **Reads Downsampled** | **Genome Coverage, % [Total/P)]^&^** | **Insert Size, bp**  **(P25/Median/P75)** |
| --- | --- | --- | --- | --- | --- | --- |
| 608 | Urine | 12.8 | 8.90E7 | 8.90E7 | 60.6 / 88.7 | 149 / 181 / 242 |
|  | Plasma | 43.7 | 1.69E8 | 8.90E7 | 53.5 / 100 | 161 / 171 / 186 |
| 649 | Urine | 22.4 | 1.47E8 | 1.47E8 | 64.6 / 87.5 | 148 / 179 / 233 |
|  |  |  | 5.90E8^#^ | 1.47E8 | 80.8 / 100 | 126 / 174 / 255 |
|  | Plasma | 44.3 | 2.05E8 | 1.48E8 | 66.0 / 94.4 | 163 / 174 / 190 |
|  |  |  | 5.45E8^#^ | 1.48E8 | 77.6 / 100 | 162 / 174 / 193 |
| 697 | Urine | 10.0 | 1.04E8 | 1.04E8 | 64.6 / 87.6 | 137 / 172 / 216 |
|  | plasma | 78.3 | 2.46E8 | 1.04E8 | 64.0 / 100 | 161 / 172 / 188 |

* DNA amount used for library preparation

\# Data obtained from the second, deeper NGS run

& Overall genome fraction covered (total) / urine-covered positions as a fraction of plasma-covered positions (P)

### **Table S6.** Mutation detection by qPCR in plasma and urine of patients with HCC (n=101).

| **Pt ID** | **Age** | **Gender** | **AFP ng/ml** | **TP53 mutation** | | **CTNNB1 mutation** | | **Tert mutation** | | **Viral etiology (HBV/HCV)** |
| --- | --- | --- | --- | --- | --- | --- | --- | --- | --- | --- |
|  |  |  |  | **Plasma** | **Urine** | **Plasma** | **Urine** | **Plasma** | **Urine** |  |
| 1 | 66-70 | F | 48,326 | BLOD | BLOD | BLOD | BLOD | BLOD | BLOD | - |
| 2 | 51-55 | M | 3 | BLOD | BLOD | BLOD | BLOD | BLOD | BLOD | - |
| 3 | 51-55 | F | 6,420 | BLOD | BLOD | BLOD | BLOD | + | BLOD | HCV |
| 4 | 31-35 | M | 1,640 | BLOD | BLOD | BLOD | BLOD | + | BLOD | - |
| 5 | 61-65 | M | 140 | BLOD | BLOD | BLOD | BLOD | + | BLOD | HCV |
| 6 | 56-60 | M | 901 | BLOD | BLOD | BLOD | BLOD | BLOD | BLOD | HBV |
| 7 | 51-55 | M | 72 | BLOD | BLOD | BLOD | BLOD | + | BLOD | HBV |
| 8 | 66-70 | M | 4 | BLOD | BLOD | BLOD | BLOD | + | BLOD | - |
| 9 | 66-70 | M | 13,522 | BLOD | BLOD | BLOD | BLOD | + | BLOD | HCV |
| 10 | 61-65 | M | 13 | BLOD | BLOD | BLOD | BLOD | + | BLOD | - |
| 11 | 51-55 | M | 38 | BLOD | BLOD | + | + | + | + | HCV |
| 12 | 51-55 | M | 58 | BLOD | BLOD | BLOD | BLOD | + | + | HBV, HCV |
| 13 | 66-70 | M | 35 | BLOD | BLOD | BLOD | BLOD | + | BLOD | HBV, HCV |
| 14 | 61-65 | M | 3.68 | BLOD | BLOD | BLOD | BLOD | BLOD | BLOD | HBV |
| 15 | 56-60 | M | 105.2 | BLOD | BLOD | BLOD | BLOD | BLOD | BLOD | HBV |
| 16 | 66-70 | M | 12.02 | BLOD | BLOD | BLOD | BLOD | BLOD | BLOD | HCV |
| 17 | 61-65 | M | 3.26 | BLOD | BLOD | BLOD | BLOD | BLOD | BLOD | HCV |
| 18 | 41-45 | M | 2.96 | BLOD | BLOD | BLOD | BLOD | BLOD | BLOD | HBV |
| 19 | 56-60 | M | 3.18 | BLOD | BLOD | BLOD | BLOD | + | + | HBV |
| 20 | 71-75 | M | 588.3 | BLOD | BLOD | BLOD | BLOD | BLOD | BLOD | HBV |
| 21 | 56-60 | M | 97.93 | BLOD | BLOD | BLOD | BLOD | BLOD | BLOD | HBV |
| 22 | 61-65 | M | 16.2 | BLOD | BLOD | BLOD | BLOD | BLOD | BLOD | HBV |
| 23 | 66-70 | M | 2.35 | BLOD | BLOD | BLOD | BLOD | BLOD | BLOD | - |
| 24 | 81-85 | F | 2.92 | BLOD | BLOD | + | BLOD | + | + | - |
| 25 | 66-70 | F | 44.8 | + | BLOD | + | BLOD | BLOD | BLOD | HCV |
| 26 | 61-65 | M | <1.3 | BLOD | BLOD | BLOD | BLOD | + | + | HBV |
| 27 | 76-80 | F | 3 | BLOD | BLOD | BLOD | + | + | + | HCV, HBV |
| 28 | 51-55 | M | 1.8 | BLOD | BLOD | BLOD | BLOD | BLOD | + | HBV |
| 29 | 61-65 | M | 2.9 | BLOD | + | BLOD | + | + | + | HBV |
| 30 | 66-70 | M | 12 | BLOD | BLOD | BLOD | BLOD | + | BLOD | HBV |
| 31 | 56-60 | M | 79.85 | BLOD | BLOD | BLOD | BLOD | BLOD | BLOD | HBV |
| 32 | 66-70 | F | 3.6 | BLOD | BLOD | BLOD | BLOD | NA | NA | HBV |
| 33 | 76-80 | F | 1.6 | BLOD | + | BLOD | BLOD | NA | NA | HBV |
| 34 | 46-50 | M | 2 | BLOD | + | + | + | NA | NA | HBV |
| 35 | 46-50 | M | 2 | BLOD | BLOD | BLOD | BLOD | NA | NA | HBV |
| 36 | 86-90 | M | 270.6 | BLOD | BLOD | BLOD | BLOD | NA | NA | - |
| 37 | 51-55 | M | 38 | BLOD | BLOD | + | BLOD | NA | NA | HCV |
| 38 | 56-60 | M | 7.64 | BLOD | BLOD | BLOD | BLOD | NA | NA | HBV |
| 39 | 56-60 | F | 22,366 | BLOD | BLOD | BLOD | BLOD | NA | NA | HBV |
| 40 | 71-75 | M | 1.5 | BLOD | BLOD | BLOD | BLOD | NA | NA | HBV |
| 41 | 66-70 | M | 105 | BLOD | + | NA | NA | BLOD | + |  |
| 42 | 61-65 | M | 3 | BLOD | BLOD | NA | NA | BLOD | + | HCV |
| 43 | 56-60 | M | 30 | BLOD | BLOD | NA | NA | + | + | HCV |
| 44 | 61-65 | M | 151 | BLOD | BLOD | NA | NA | BLOD | + | HCV |
| 45 | 66-70 | F | 577 | BLOD | BLOD | NA | NA | BLOD | + | HCV |
| 46 | 76-80 | M | 5 | BLOD | + | NA | NA | + | BLOD | - |
| 47 | 56-60 | M | 336 | BLOD | BLOD | NA | NA | + | BLOD | HCV |
| 48 | 56-60 | M | 41.9 | BLOD | + | NA | NA | NA | NA | HCV |
| 49 | 71-75 | F | 4 | BLOD | + | NA | NA | + | BLOD | HBV |
| 50 | 61-65 | M | 2 | BLOD | BLOD | NA | NA | BLOD | BLOD | HCV |
| 51 | 56-60 | M | 4.9 | BLOD | BLOD | NA | NA | NA | NA | HCV |
| 52 | 56-60 | F | 3.5 | BLOD | BLOD | NA | NA | BLOD | BLOD | HCV |
| 53 | 71-75 | F | 10.3 | BLOD | BLOD | NA | NA | BLOD | + | HBV, HCV |
| 54 | 71-75 | F | 11.8 | BLOD | BLOD | NA | NA | + | BLOD | HCV |
| 55 | 66-70 | F | 45.7 | BLOD | BLOD | NA | NA | BLOD | BLOD | - |
| 56 | 81-85 | F | 34.7 | BLOD | BLOD | NA | NA | BLOD | BLOD | HCV |
| 57 | 86-90 | M | 3.3 | + | BLOD | NA | NA | + | BLOD | HBV |
| 58 | 66-70 | M | 1.9 | BLOD | BLOD | NA | NA | BLOD | BLOD | HBV |
| 59 | 56-60 | F | 591.4 | BLOD | BLOD | NA | NA | + | BLOD | HCV |
| 60 | 51-55 | M | 167.4 | BLOD | BLOD | NA | NA | BLOD | BLOD | HBV |
| 61 | 76-80 | M | n/a | BLOD | BLOD | NA | NA | BLOD | BLOD | - |
| 62 | 76-80 | M | 5 | BLOD | BLOD | NA | NA | BLOD | BLOD | HBV |
| 63 | 66-70 | M | 1264 | BLOD | BLOD | NA | NA | BLOD | BLOD | - |
| 64 | 51-55 | M | 5 | BLOD | BLOD | NA | NA | BLOD | BLOD | HBV |
| 65 | 61-65 | M | 7.5 | BLOD | BLOD | NA | NA | BLOD | BLOD | HBV |
| 66 | 76-80 | F | 833.6 | BLOD | BLOD | NA | NA | BLOD | BLOD | HCV |
| 67 | 71-75 | M | 2.8 | BLOD | BLOD | NA | NA | BLOD | BLOD | HCV |
| 68 | 61-65 | F | n/a | BLOD | BLOD | NA | NA | BLOD | BLOD | HBV |
| 69 | 81-85 | F | n/a | BLOD | BLOD | NA | NA | BLOD | BLOD | HBV |
| 70 | 76-80 | M | 12,179 | BLOD | BLOD | NA | NA | NA | NA | - |
| 71 | 66-70 | M | 6.7 | BLOD | + | NA | NA | NA | NA | HBV |
| 72 | 76-80 | F | 14.2 | BLOD | BLOD | NA | NA | NA | NA | HCV |
| 73 | 61-65 | M | 9.6 | BLOD | BLOD | NA | NA | NA | NA | HBV |
| 74 | 66-70 | M | n/a | BLOD | BLOD | NA | NA | NA | NA | HBV |
| 75 | 66-70 | M | n/a | BLOD | BLOD | NA | NA | NA | NA | HBV |
| 76 | 61-65 | F | n/a | BLOD | BLOD | NA | NA | NA | NA | HBV |
| 77 | 66-70 | F | n/a | BLOD | BLOD | NA | NA | NA | NA | HCV |
| 78 | 61-65 | F | n/a | BLOD | BLOD | NA | NA | NA | NA | HBV |
| 79 | 76-80 | F | n/a | BLOD | BLOD | NA | NA | NA | NA | HBV |
| 80 | 61-65 | M | n/a | BLOD | BLOD | NA | NA | NA | NA | HBV |
| 81 | 51-55 | M | 18,733 | BLOD | BLOD | NA | NA | BLOD | + | HBV, HCV |
| 82 | 61-65 | M | 7.3 | NA | NA | BLOD | BLOD | BLOD | BLOD | HBV |
| 83 | 66-70 | F | 430 | NA | NA | BLOD | BLOD | + | + | HBV, HCV |
| 84 | 31-35 | M | 4861 | NA | NA | BLOD | BLOD | + | + | HBV |
| 85 | 61-65 | F | 4.57 | NA | NA | BLOD | BLOD | BLOD | BLOD | HBV |
| 86 | 56-60 | M | 17 | NA | NA | BLOD | + | NA | NA | HCV |
| 87 | 76-80 | M | 1,596 | NA | NA | + | BLOD | NA | NA | - |
| 88 | 61-65 | M | <1.3 | NA | NA | BLOD | BLOD | NA | NA | HBV |
| 89 | 61-65 | M | 3.42 | NA | NA | + | BLOD | NA | NA | HBV |
| 90 | 26-30 | M | 3 | NA | NA | NA | NA | BLOD | + | - |
| 91 | 66-70 | M | 104.5 | NA | NA | NA | NA | BLOD | + | - |
| 92 | 66-70 | F | 12,983 | NA | NA | NA | NA | + | + | - |
| 93 | 61-65 | F | 75,917 | NA | NA | NA | NA | + | + | HBV |
| 94 | 71-75 | F | 836,322 | NA | NA | NA | NA | BLOD | BLOD | HBV |
| 95 | 56-60 | F | 4,489 | NA | NA | NA | NA | + | BLOD | HCV |
| 96 | 66-70 | M | 1,103 | NA | NA | NA | NA | BLOD | BLOD | - |
| 97 | 51-55 | M | 595 | NA | NA | NA | NA | BLOD | BLOD | HCV |
| 98 | 71-75 | F | n/a | NA | NA | NA | NA | BLOD | BLOD | HCV |
| 99 | 61-65 | M | 11.4 | NA | NA | NA | NA | BLOD | BLOD | - |
| 100 | 61-65 | M | 8.2 | NA | NA | NA | NA | BLOD | BLOD | HBV |
| 101 | 71-75 | M | 361 | NA | NA | NA | NA | + | BLOD | HBV |

BLOD, below limit of detection; highlighted for positive for detection; black cells, test not performed due to limited quantity of DNA .

### **Table S7.** Mutations detected by qPCR in urine-plasma pairs with at least one positive test (n, number of pairs).

|  | Plasma | Both | Urine | p value *  (plasma *vs.* urine) |
| --- | --- | --- | --- | --- |
| TP53 (n=10) | 2 | 0 | 8 | 0.023 |
| CTNNB1 (n=10) | 7 | 2 | 5 | 0.65 |
| TERT (n=39) | 29 | 12 | 22 | 0.475 |

**Fisher’s Exact Test*

### **Table S8.** Targeted NGS summary.

| Patient ID | Mean Unique Coverage of Target Positions* | | Mean Variant Call Rate Across All Targets** | |
| --- | --- | --- | --- | --- |
|  | Plasma | Urine | Plasma | Urine |
| HCC-008 | 1,277 | 1,064 | 0.0182 | 0.0207 |
| HCC-030 | 1,167 | 1,421 | 0.0153 | 0.0204 |
| HCC-054 | 924 | 262 | 0.0131 | 0.0092 |
| HCC-057 | 1,565 | 546 | 0.0229 | 0.0128 |
| HCC-058 | 855 | 1,251 | 0.0122 | 0.0233 |
| HCC-060 | 1,245 | 2,089 | 0.0145 | 0.0322 |
| HCC-063 | 657 | 985 | 0.0123 | 0.0179 |
| HCC-068 | 2,440 | 1,391 | 0.0288 | 0.0327 |
| HCC-069 | 3,872 | 859 | 0.0379 | 0.0205 |
| HCC-079 | 340 | 2,852 | 0.0105 | 0.1027 |
| HCC-277 | 795 | 1,637 | 0.0133 | 0.0312 |
| HCC-297 | 1,206 | 1,660 | 0.0189 | 0.0198 |
| HCC-326 | 3,044 | 1,942 | 0.0219 | 0.0290 |
| HCC-335 | 1,159 | 899 | 0.0125 | 0.0175 |
| HCC-724 | 1,957 | 286 | 0.0265 | 0.0085 |

* UMI family size ≥3
** Variant calls supported by ≥3 UMI families

### **Table S9.** Global concordance by position (≥3 supporting UMI families).

| **Position Call** | | **Plasma** | **Urine** |
| --- | --- | --- | --- |
| negative (reference) | | 942,253 | 934,576 |
| positive (variant) | | 17,837 | 25,514 |
| total | | 960,090 | 960,090 |
| positive concordant | | 6,506 | |
| negative concordant | | 923,245 | |
| **Concordance, %** | overall^a^ | 96.8 | |
|  | positive^b^ | 17.7 | |
|  | plasma-positive^c^ | 36.5 | |
|  | urine-positive^d^ | 25.5 | |

^a.^ percentage of positive and negative concordants over total positions (6,506 + 923,245/960,090)

^b.^ percentage of positive concordants over total positive variants (6,506/17,837 + 24,514 - 6,506)

^c.^ percentage of positive concordants over plasma variants (6,506/17,837)

^d.^ percentage of positive concordants over urine variants (6,506/25,514)

### **Table S10.** Somatic (VAF<0.4) variant call rates by target in the 15-patient HCC cohort (≥3 supporting UMI families).

| **Target Gene** | **Plasma** | **Urine** |
| --- | --- | --- |
| TERT promoter | 0.2023 | 0.2189 |
| KMT2D | 0.0367 | 0.0468 |
| TP53 | 0.0275 | 0.0348 |
| ARID1A | 0.0242 | 0.0355 |
| KEAP1 | 0.0232 | 0.0320 |
| AXIN1 | 0.0199 | 0.0276 |
| PCLO | 0.0199 | 0.0250 |
| CDKN2A | 0.0121 | 0.0259 |
| NFE2L2 | 0.0193 | 0.0185 |
| TSC2 | 0.0120 | 0.0221 |
| HNF1A | 0.0113 | 0.0226 |
| ARID1B | 0.0098 | 0.0210 |
| ALB | 0.0119 | 0.0170 |
| APC | 0.0111 | 0.0164 |
| LRP1B | 0.0108 | 0.0166 |
| APOB | 0.0087 | 0.0168 |
| BAP1 | 0.0075 | 0.0169 |
| ATM | 0.0101 | 0.0138 |
| ARID2 | 0.0075 | 0.0145 |
| CTNNB1 | 0.0093 | 0.0122 |
| RB1 | 0.0085 | 0.0128 |
| IL6ST | 0.0056 | 0.0124 |
| PIK3CA | 0.0051 | 0.0122 |
| RPS6KA3 | 0.0070 | 0.0070 |

### **Table S11**. Comparison of genome coverage by PBMC and total urine DNA.

| **ID** | **DNA** | **# Reads** | **Depth** | **% Aligned**  **(Read 1/2)** | **% Q30 (Read 1/2)** | **% Genome** | **% PBMC** |
| --- | --- | --- | --- | --- | --- | --- | --- |
| 1 | Urine | 864,623,648 | 39.23 | 99.15 (49.22 / 49.93) | 92.43 (46.43 / 46.00) | 91.68 | 99.90 |
|  | PBMC | 700,019,656 | 32.39 | 98.94 (50.32 / 48.63) | 93.59 (47.81 / 45.77) | 91.35 | 100 |
| 2 | Urine | 899,433,124 | 41.01 | 99.45 (49.65 / 49.80) | 94.00 (47.12 / 46.88) | 91.02 | 99.28 |
|  | PBMC | 701,363,735 | 32.14 | 99.57 (49.53 / 50.04) | 93.31 (46.68 / 46.64) | 91.09 | 100 |
| 3 | Urine | 1,162,778,516 | 52.66 | 98.08 (48.62 / 49.47) | 92.21 (46.32 / 45.89) | 92.24 | 99.94 |
|  | PBMC | 939,328,559 | 43.12 | 99.44 (49.56 / 49.88) | 93.28 (46.65 / 46.63) | 91.31 | 100 |
| 4 | Urine | 819,973,434 | 37.43 | 99.64 (49.59 / 50.05) | 92.57 (46.32 / 46.24) | 92.14 | 99.76 |
|  | PBMC | 796,160,774 | 36.85 | 99.51 (49.59 / 49.92) | 93.05 (46.56 / 46.49) | 92.03 | 100 |
| 5 | Urine | 550,159,977 | 24.26 | 96.21 (47.93 / 48.29) | 92.08 (46.11 / 45.98) | 92.03 | 99.66 |
|  | PBMC | 811,763,599 | 37.06 | 98.25 (48.84 / 49.41) | 92.80 (46.36 / 46.44) | 91.94 | 100 |
| 6 | Urine | 896,851,243 | 40.83 | 98.60 (49.00 / 49.60) | 92.51 (46.22 / 46.28) | 91.71 | 99.41 |
|  | PBMC | 832,059,596 | 38.50 | 99.26 (49.38 / 49.88) | 93.47 (46.68 / 46.79) | 91.85 | 100 |

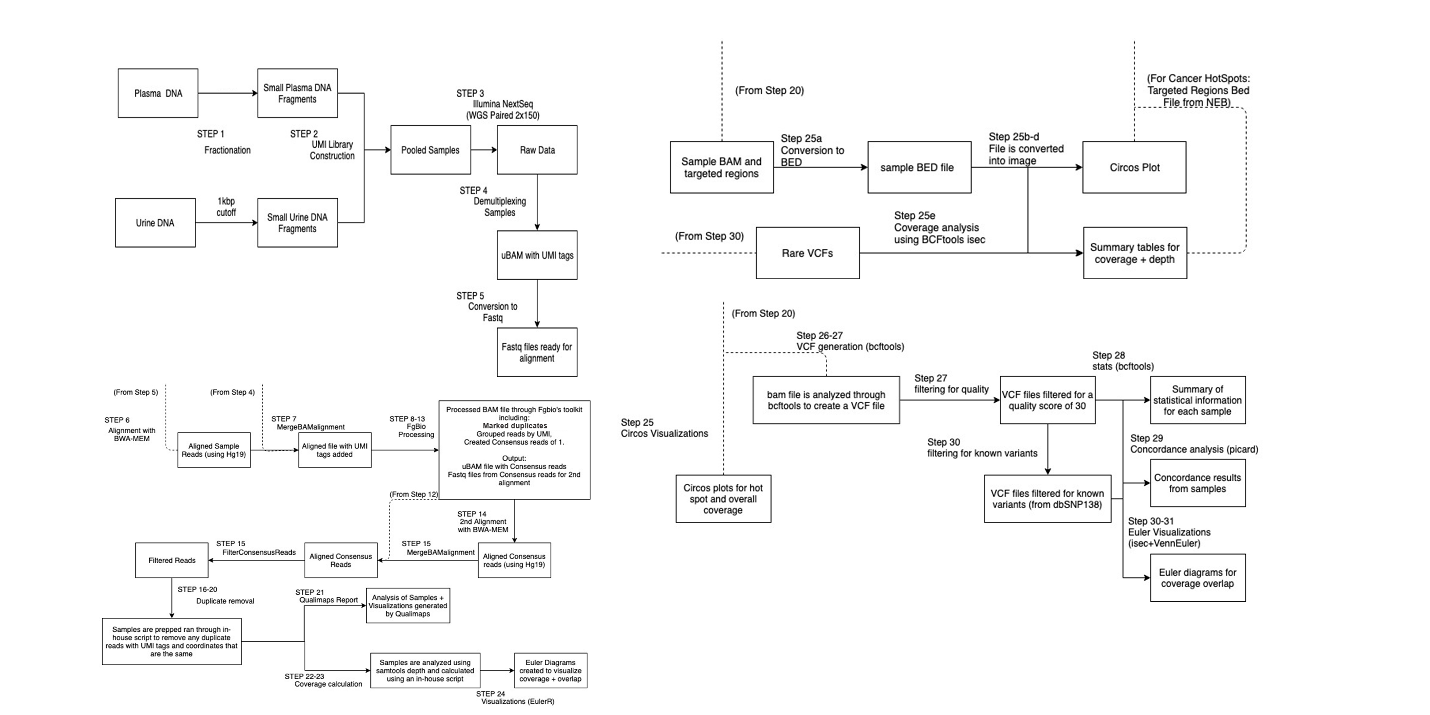

**Figure S1**. NGS data analysis workflows.

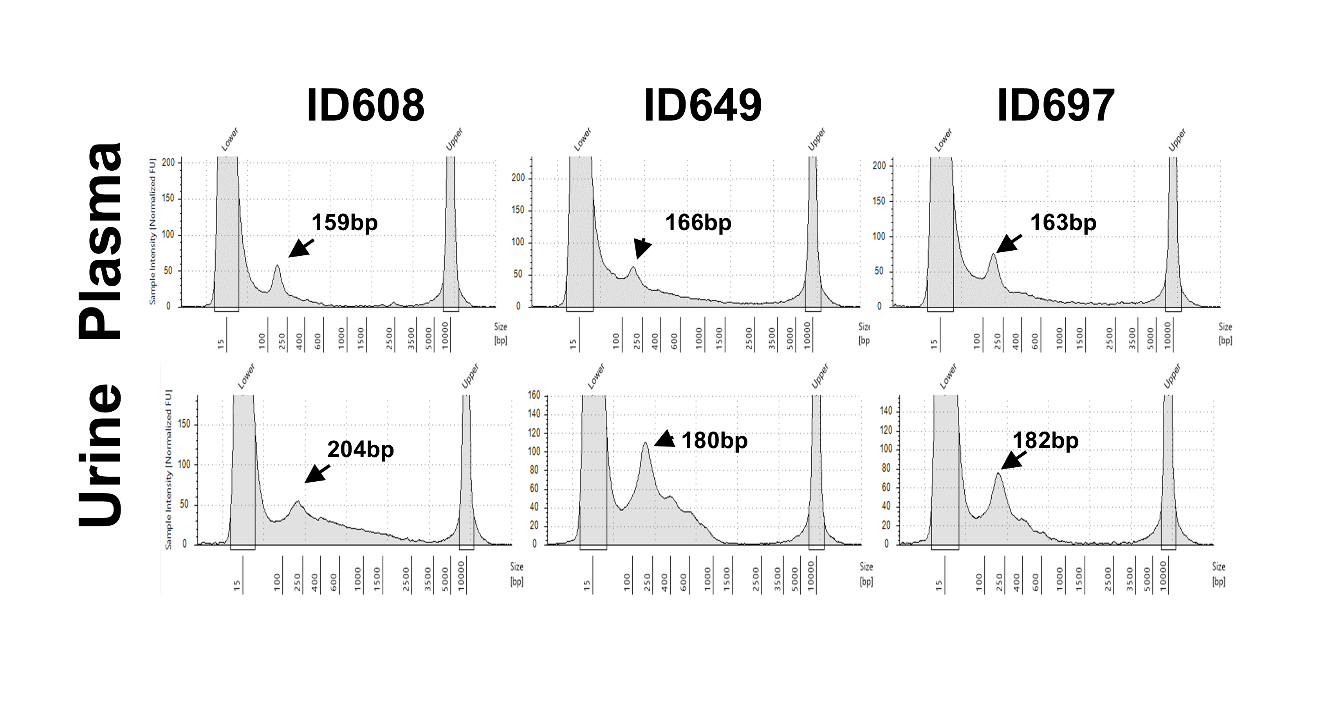

**Figure S2**. Electrophoretic size profiling of LMW DNA isolated from urine and plasma of two HCC patents.

Matched plasma and urine cfDNA are shown with DNA input equivalents of 0.33 ml and 0.0175 ml of urine or plasma, respectively. Plasma DNA was size-selected (<500bp) prior to analysis. Samples 649 and 697 were collected from the same patient 8 months apart (Table S1).

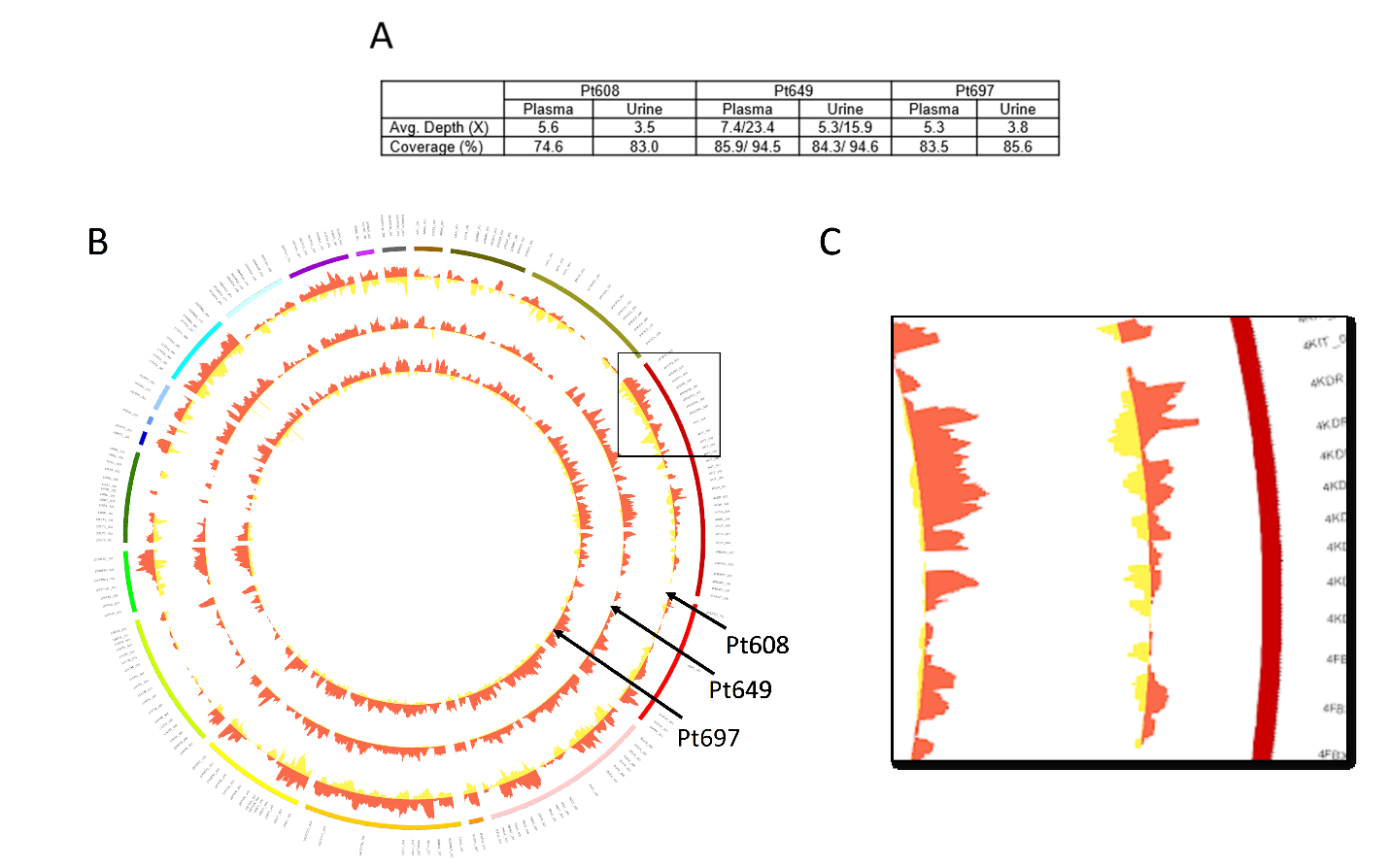

**Figure S3**. Comparison of cancer hotspot coverage between matched urine and plasma cfDNA from patients with HCC. (A) Summary of sWGS hotspot coverage in three matched urine/plasma pairs. In samples 649, the second number represents data from the deep-sequencing run. (B) Circos plot depicting the comparison of shallow WGS hotspot coverage profiles of three matched plasma (red) and urine (yellow) cfDNA sample pairs. (C) Zoom-in of the area in the rectangle.

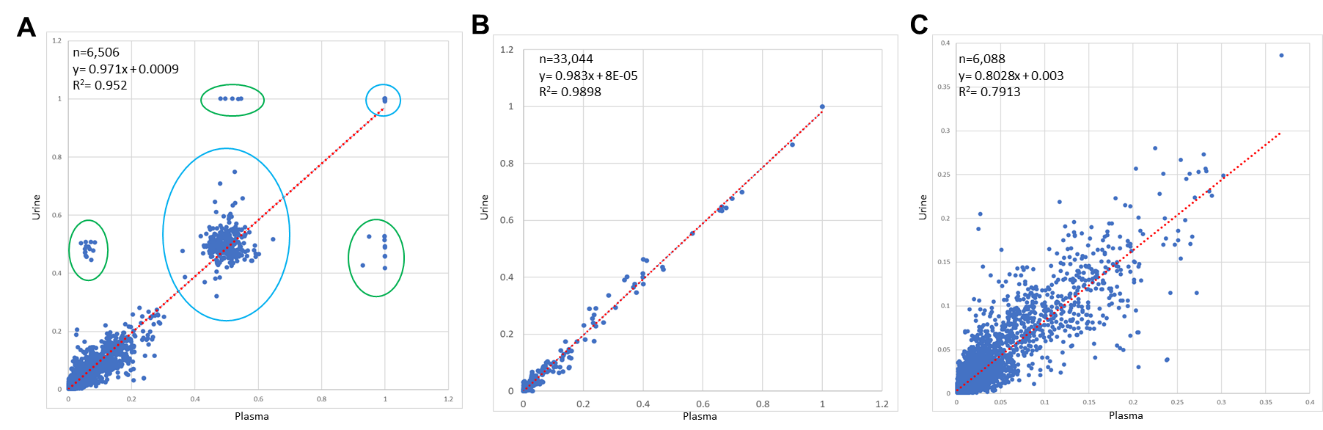

**Figure S4.** Correlation of all (A), cohort mean (B), and likely somatic (C) variant allele frequencies detected by targeted NGS in plasma or urine cfDNA of patients with HCC. Green circles indicate variants likely attributable to early embryonic somatic mutations and originating from circulating or uroepithelial cell gDNA. Blue circles indicate likely germline variants. Only variants supported by ≥3 unique reads (UMI families) were included in the analyses. n, number of double-positive positions.
