## Supplementary figures and images for "Urinary cell-free DNA as a noninvasive liquid biopsy for hepatocellular carcinoma: a novel tool for diagnostics and monitoring"

### Supplemental File 1

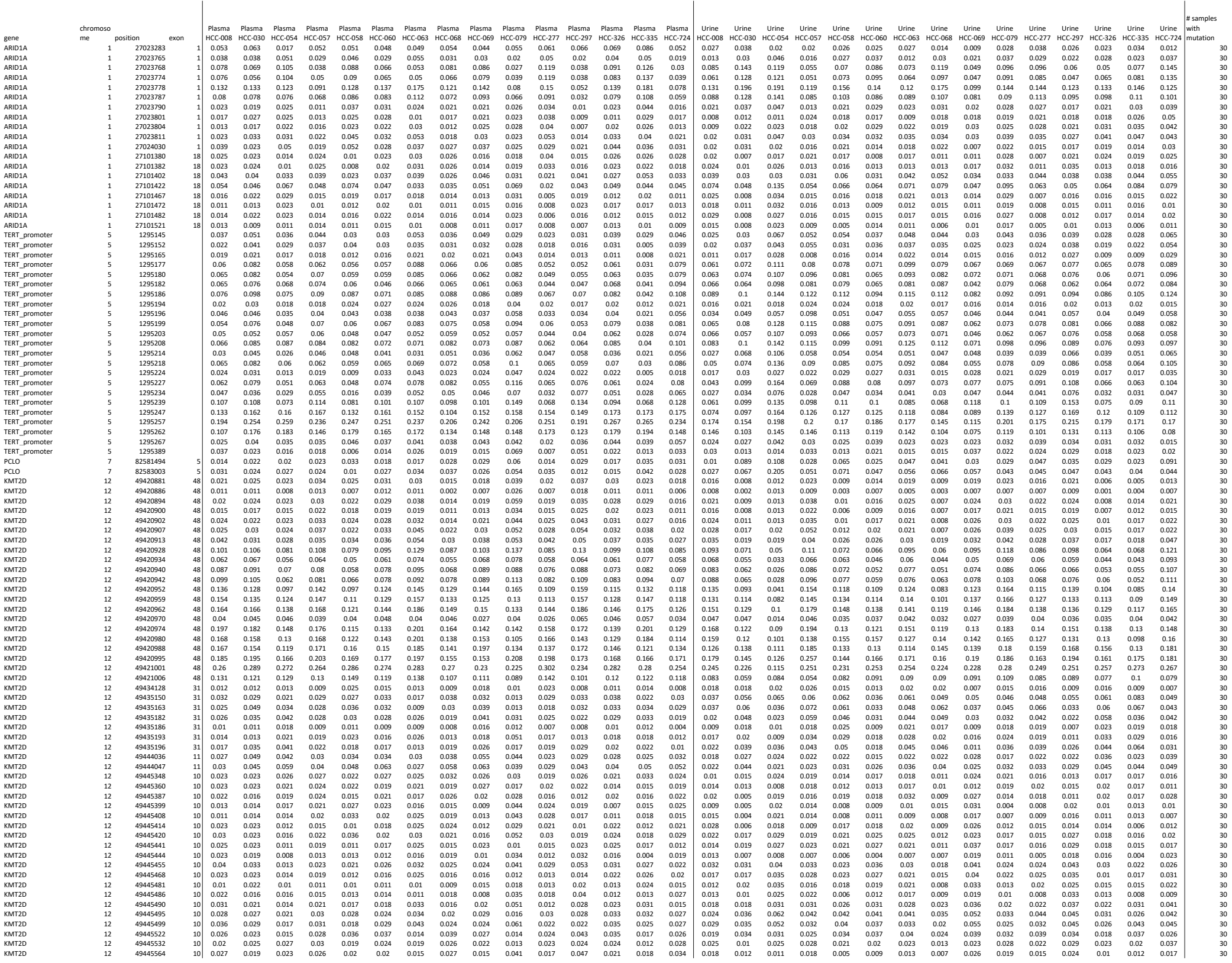

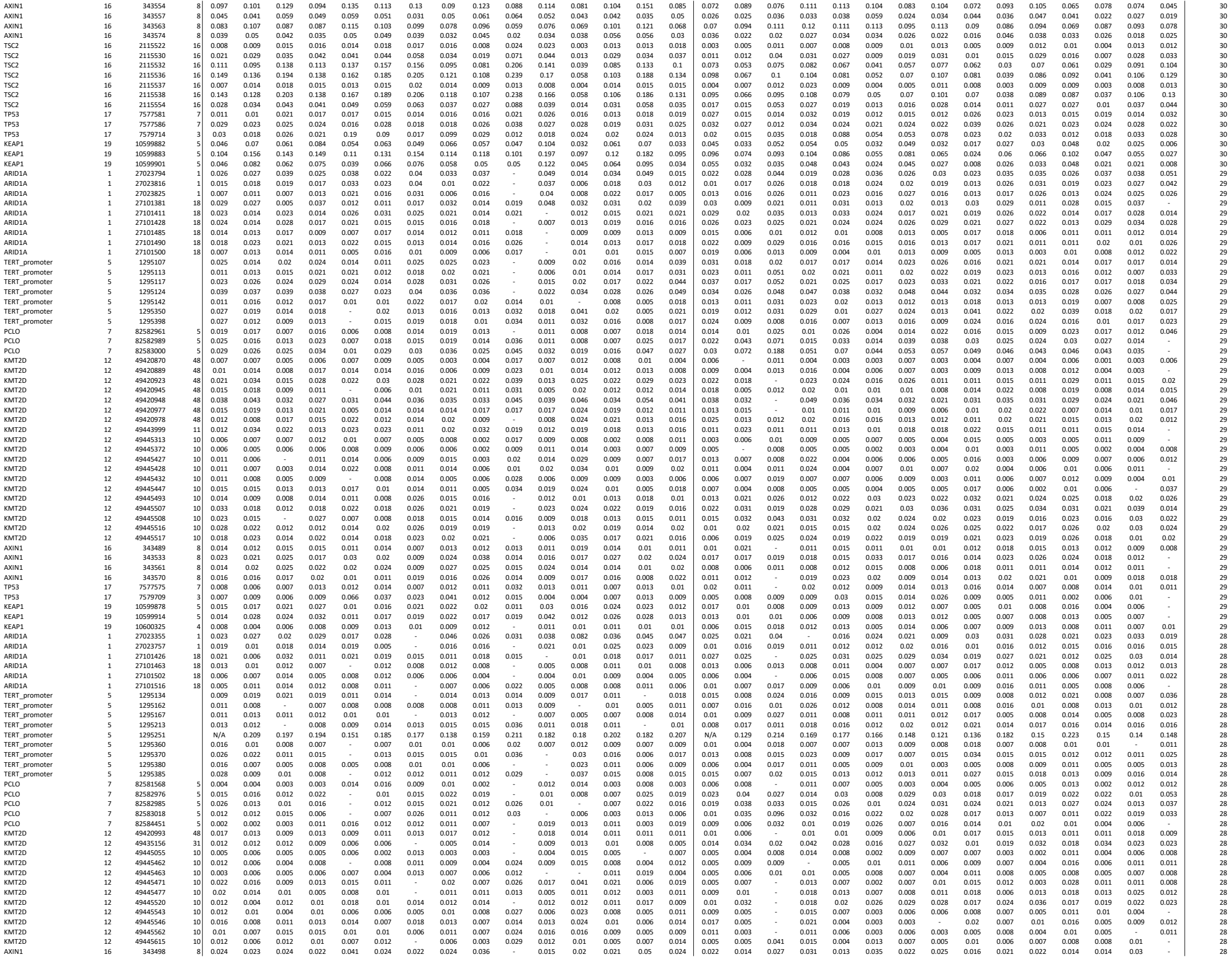

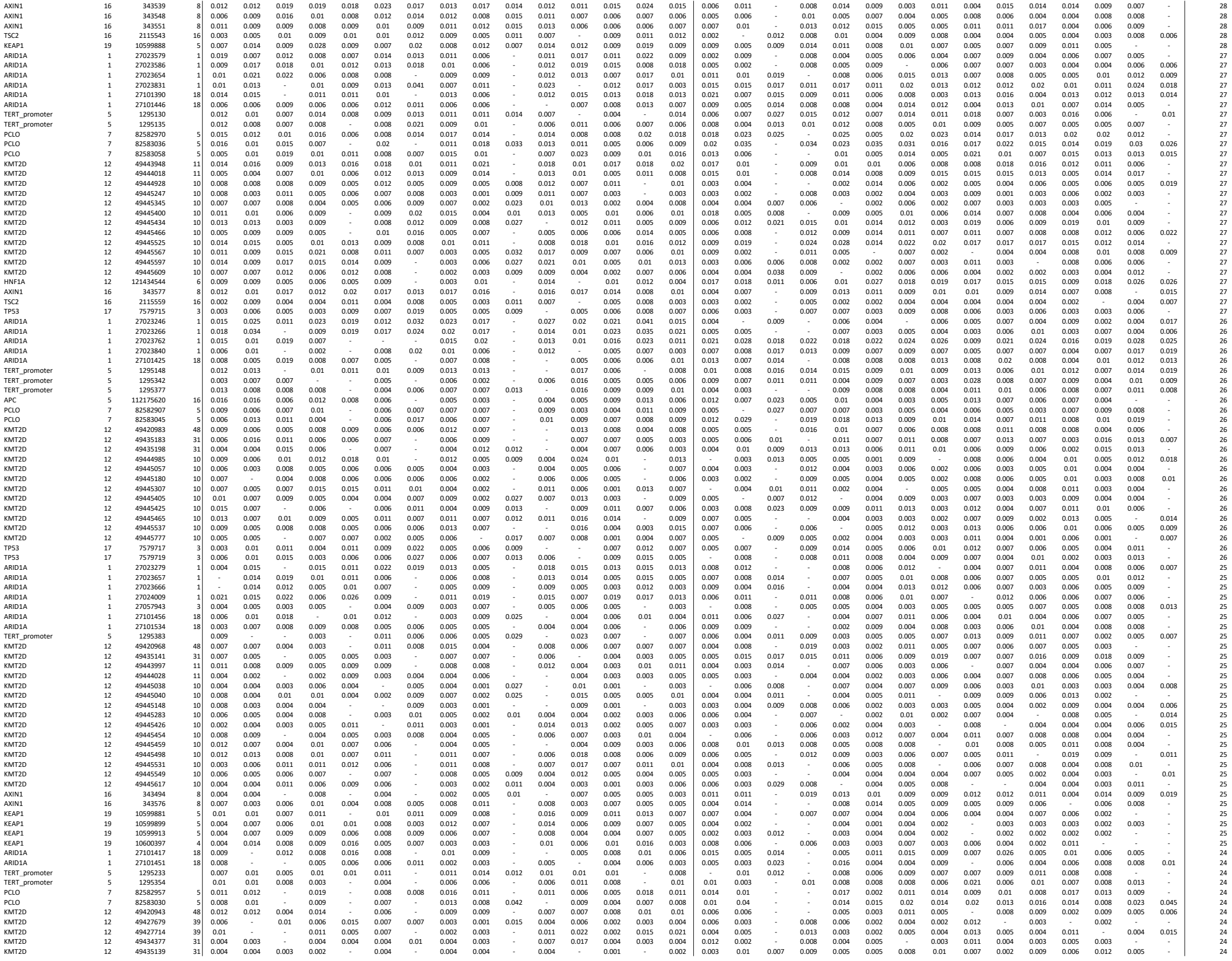

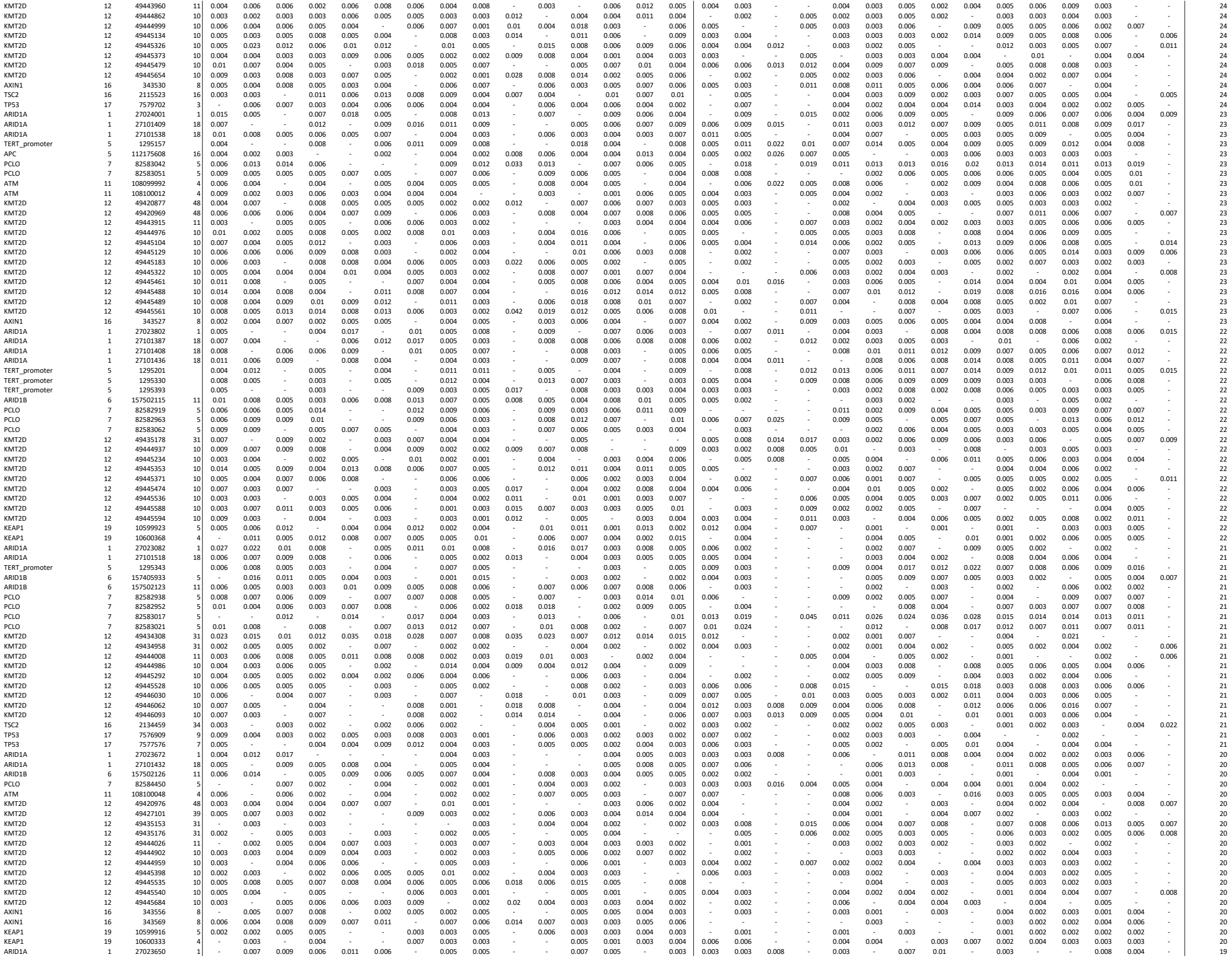

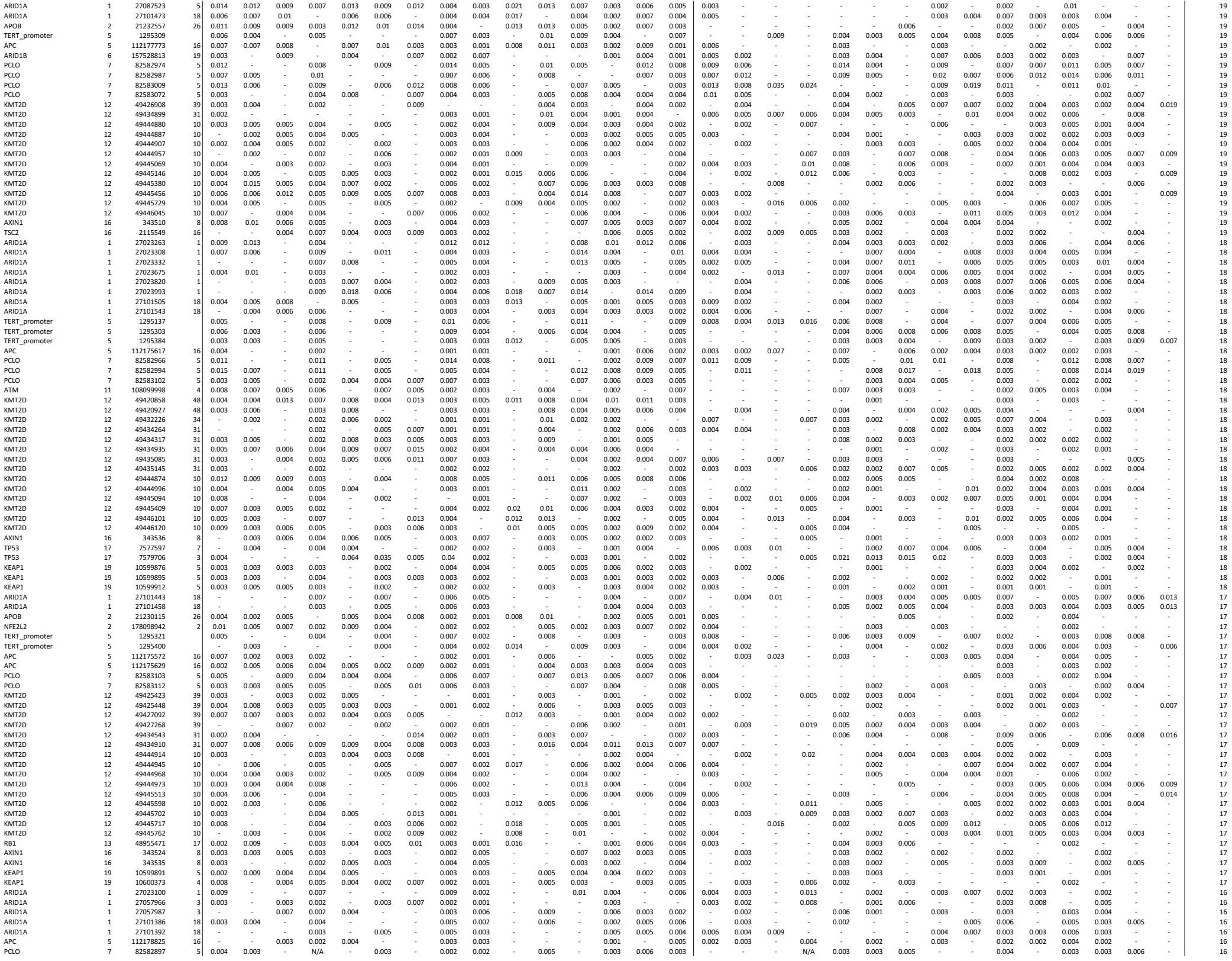

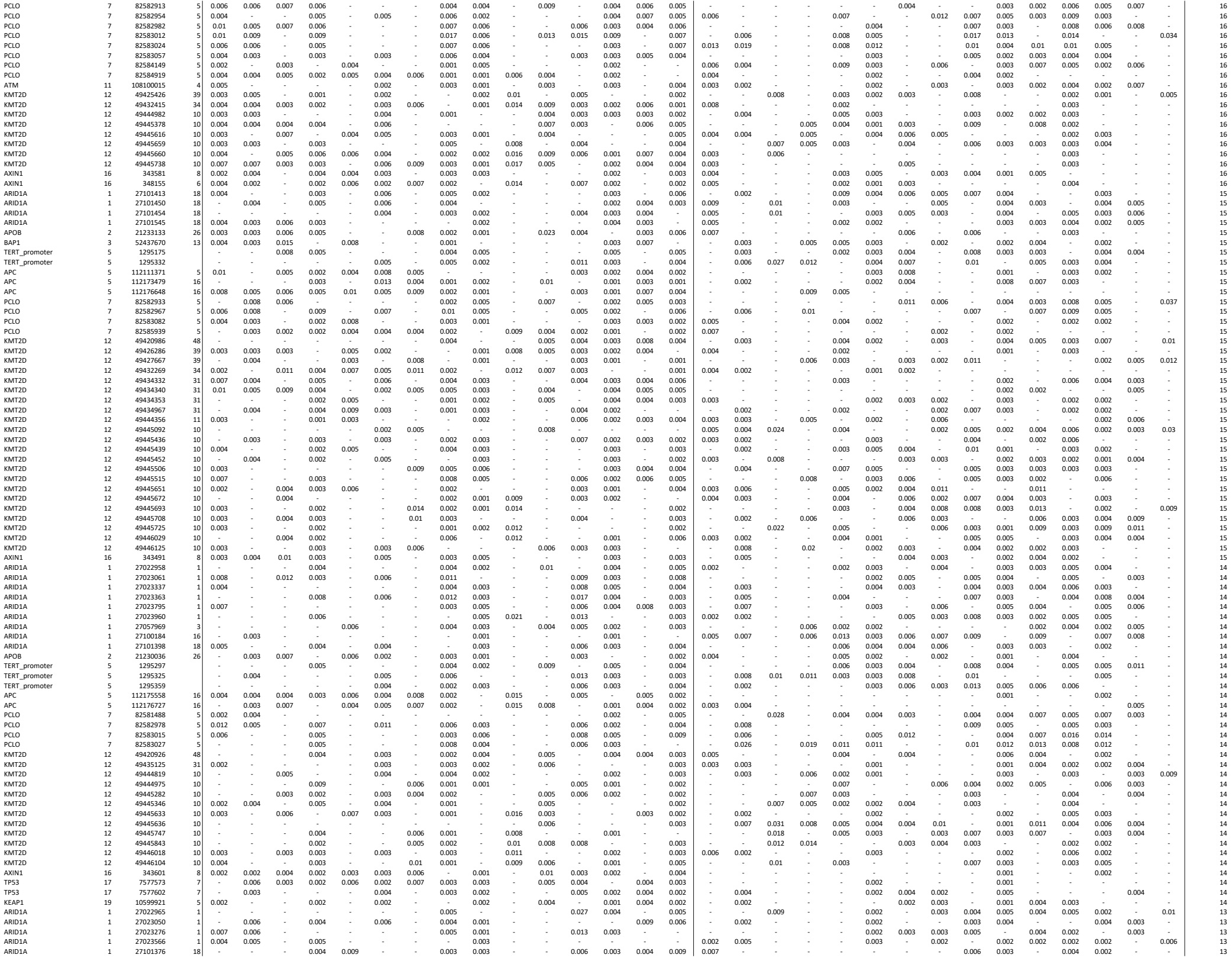

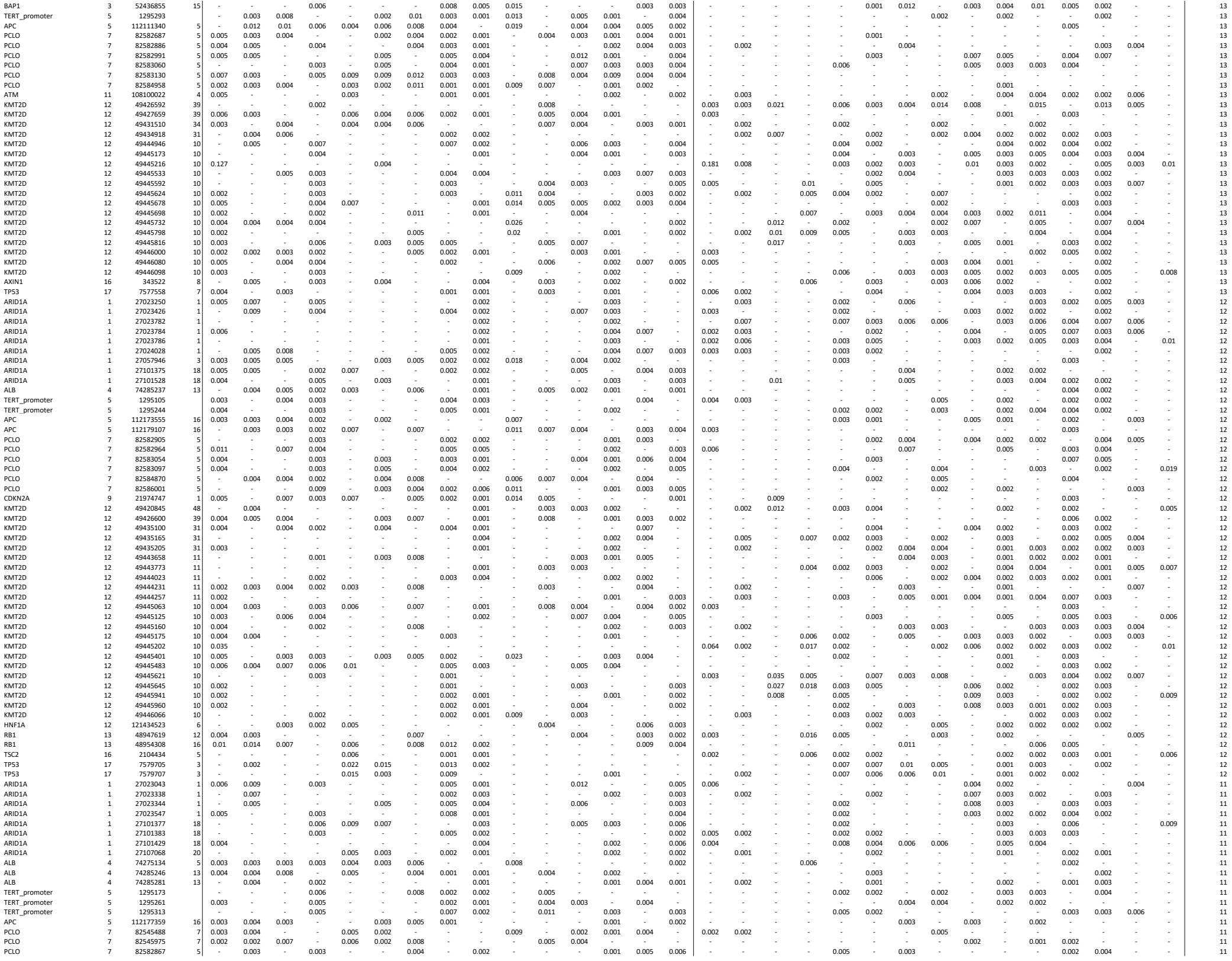

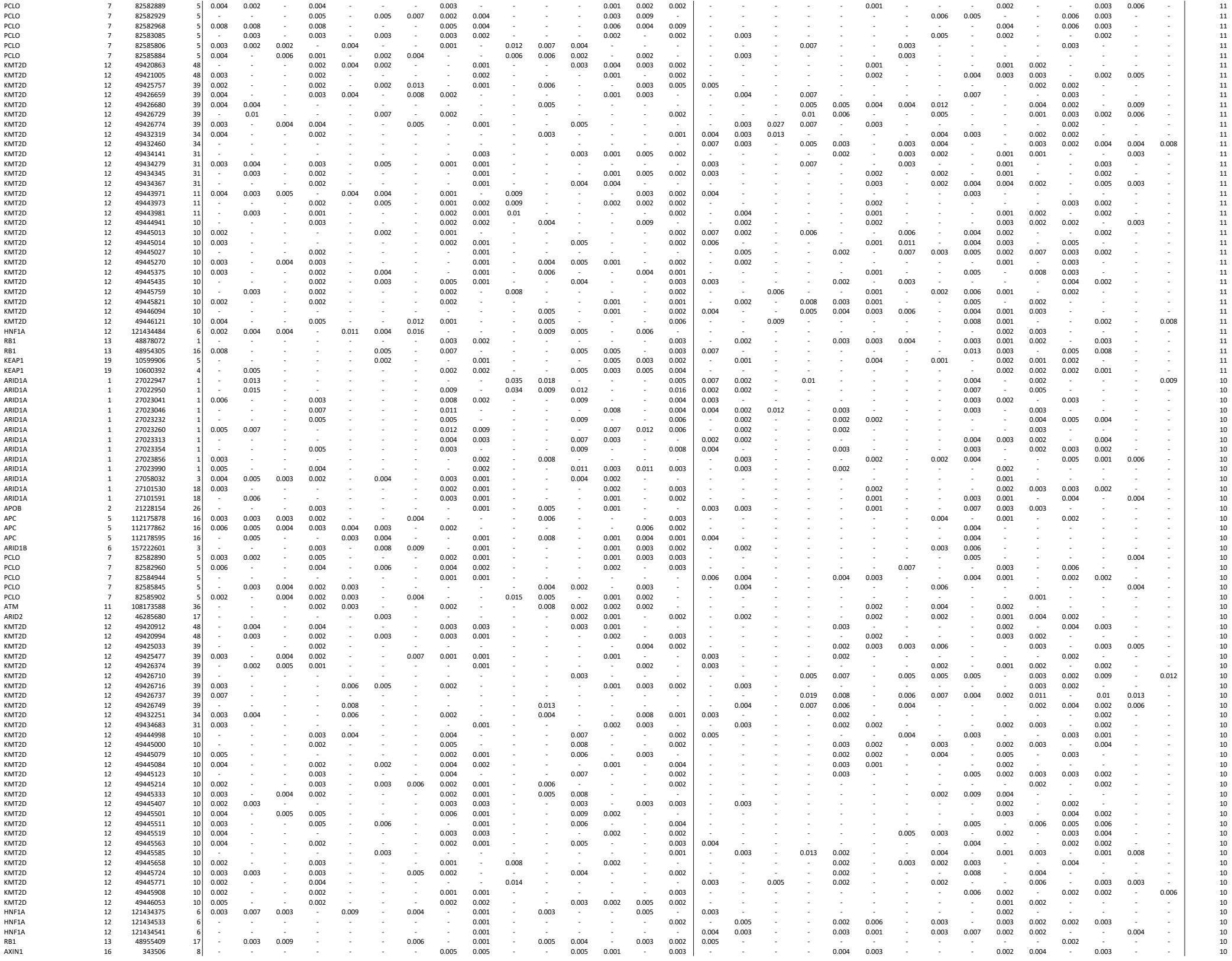

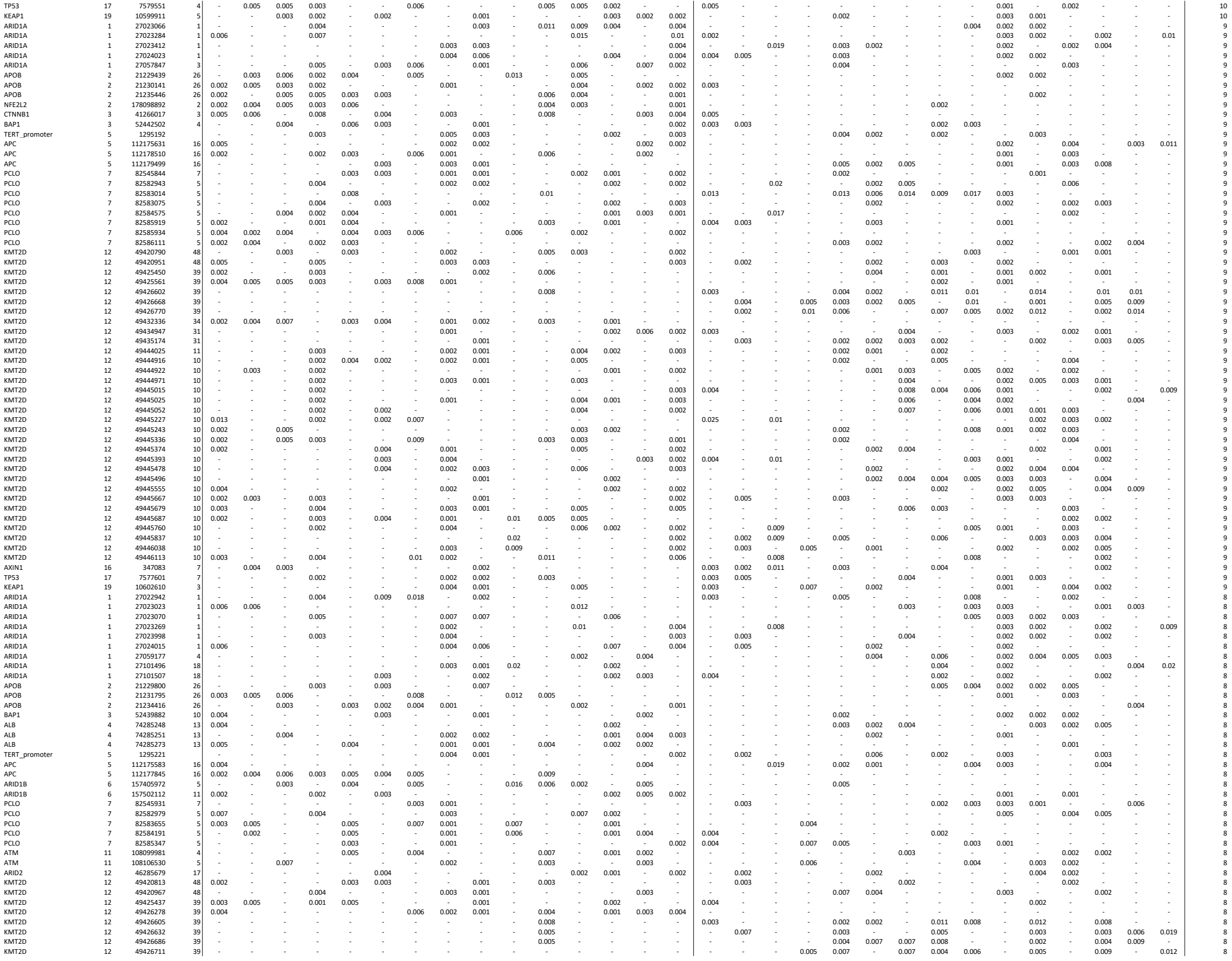

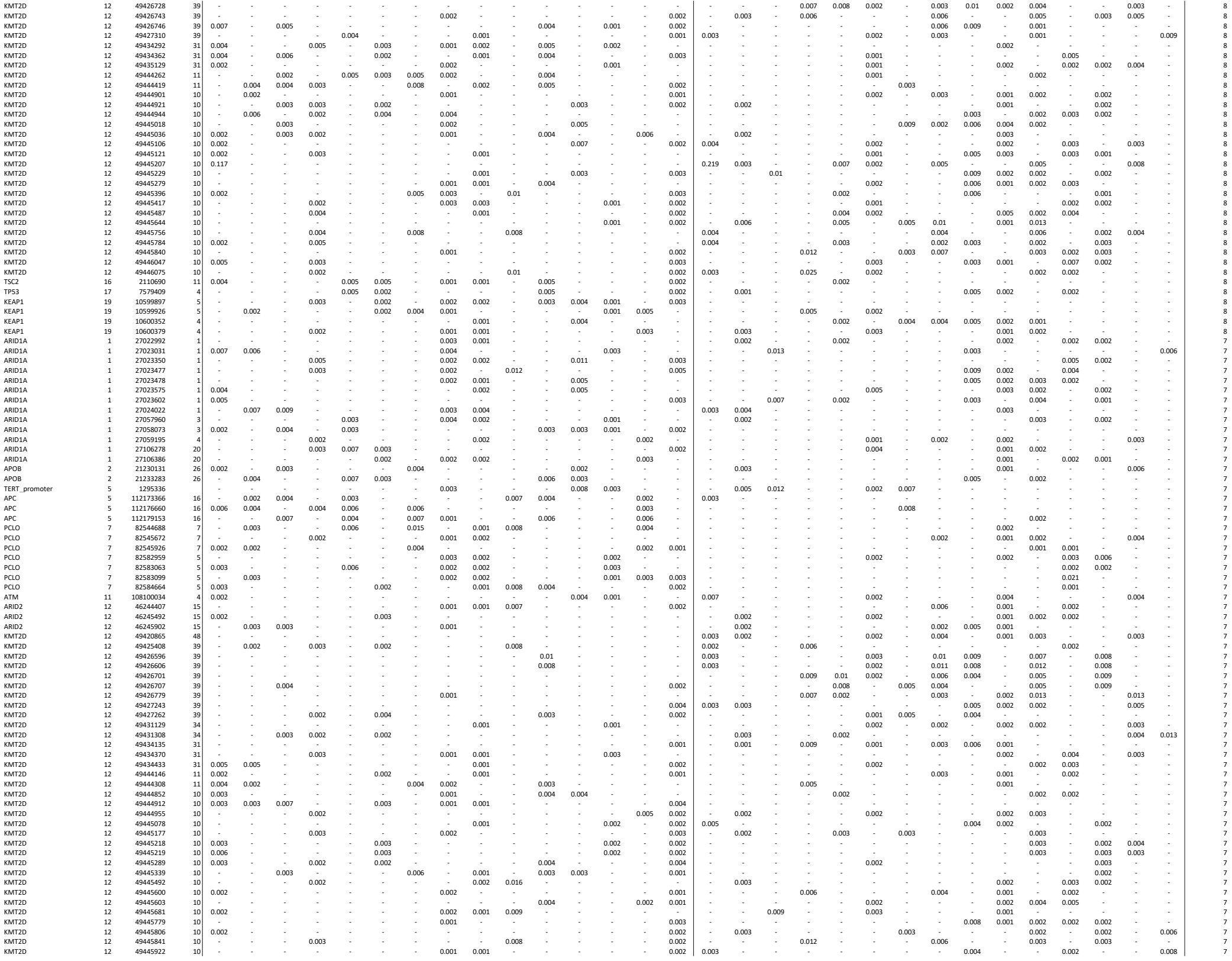
